# Hybrid novice–AI system achieves expert-level performance in intraoperative ischemia detection

**DOI:** 10.64898/2026.08.01.26359457

**Authors:** Nihal Murali, Amir I. Mina, Harsh Sinha, Joshua W. Anderson, Yash Raka, Hung-Ching Chang, Homa K. Amiri, Parthasarathy D. Thirumala, Kayhan Batmanghelich, Shyam Visweswaran

## Abstract

Carotid endarterectomy carries the risk of intraoperative cerebral ischemia, which is monitored by expert neurophysiologists through continuous electroencephalography (cEEG). Because expert availability is limited, we developed a hybrid novice–artificial intelligence (AI) system that detects ischemia using novice monitors with limited cEEG training. The hybrid system dynamically weights novice and AI inputs to arrive at a final output. Using four novices, we compared hybrid systems against experts alone, novices alone, and AI alone. Hybrid systems were statistically non-inferior to experts in sensitivity and false-positive rate (FPR), whereas novices alone were not. At 80% sensitivity, hybrid systems reduced FPR by half compared with the AI-only system, with similar benefits at 90% sensitivity. Further, the area under the precision-recall curve improved from 0.546 to 0.610–0.726, the area under the receiver operating curve improved from 0.957 to 0.967–0.971, and calibration improved compared with AI alone. These results highlight the potential of a hybrid system to monitor intraoperative cerebral ischemia.

## INTRODUCTION

Carotid endarterectomy (CEA) is performed to reduce the risk of stroke in patients with high-grade carotid stenosis, but it carries the intraoperative risk of cerebral ischemia (1,2). Intraoperative neuromonitoring (IONM) with continuous electroencephalography (cEEG) provides a reliable means of detecting ischemic changes in real time and guiding corrective actions such as shunt placement, making it the standard of care in CEA. Expert neurophysiologists with board-certified training typically perform IONM. However, due to a shortage of expert neurophysiologists, IONM is not always readily available. In the United States (U.S.), only about 4,000 certified neurophysiologists are available to cover over 750,000 procedures annually (3), a mismatch exacerbated by limited training opportunities, stringent certification requirements, and significant geographic disparities. Hospitals, therefore, experience delays or cancellations in performing CEA and struggle to provide consistent IONM coverage. These ongoing challenges highlight the unsustainability of expert-only monitoring and underscore the need for scalable solutions to expand reliable IONM capability beyond the current limitations.

Artificial intelligence (AI), including machine learning (ML) applications, has expanded rapidly in surgery, from preoperative risk stratification to intraoperative guidance and postoperative prediction (4–6). ML has also been applied to cEEG signals for seizure detection, sleep stage classification, and the diagnosis of neurological diseases such as Alzheimer’s disease, Parkinson’s disease, depression, and autism; however, applications to cerebral ischemia remain limited. While cEEG-based ML has been studied for stroke detection, these approaches typically use non-operative data that differ fundamentally from intraoperative data (7–9). In the context of CEA, prior work has used quantitative EEG (qEEG) markers of ischemia, such as spectral power, to characterize EEG changes and correlate them with ischemia (10,11). Mina et al. (12) applied supervised ML, including tree-based models such as random forests, to detect intraoperative cerebral ischemia during CEA. While these studies demonstrate the feasibility of ML-based ischemia detection, a more clinically acceptable approach is a human-plus-AI approach that augments, rather than replaces, the neurophysiologist.

In clinical practice, relying solely on either humans or AI for IONM poses significant limitations. Human-only monitoring is constrained by the scarcity of expert neurophysiologists and the cognitive demands of continuous cEEG interpretation, which can lead to fatigue-related errors. Conversely, AI models used alone remain insufficiently reliable for IONM because they exhibit overconfidence and are susceptible to spurious correlations (13–15). These complementary limitations motivate a hybrid approach that integrates human and AI strengths. In this article, we describe a hybrid novice–AI system for IONM that combines predictions from an AI model with assessments from a novice intraoperative monitor. We use the term *novice* to refer to individuals without board certification or extensive clinical experience in interpreting cEEG for intraoperative monitoring. We show that this hybrid system achieves performance that is statistically non-inferior to that of expert neurophysiologists, while achieving lower false-positive rates and higher precisions at clinically relevant sensitivities than either a novice or an AI system alone. By augmenting novice monitors without requiring expert monitoring, our approach offers a potential pathway to rapidly expand the IONM workforce.

## RESULTS

### Study design and evaluation overview

We evaluated the hybrid novice–AI system using two intraoperative cEEG datasets from patients undergoing CEA. Each cEEG recording consisted of eight channels, with four from the right hemisphere and four from the left hemisphere. From each recording, we extracted 10 minutes of cEEG signals following carotid artery clamp application, the period of highest ischemic risk. Dataset-1 included 400 patients, of whom 28 had intraoperative ischemic changes, as determined from the neurophysiologist’s intraoperative notes. Dataset-2 included 30 patients, 15 of whom exhibited ischemic changes, as determined by five expert neurophysiologists independently reviewing. Four novices with limited training in cEEG interpretation also labeled both datasets for ischemia.

These 10-minute recordings were segmented into nonoverlapping 20-second intervals, from which 111 quantitative cEEG features were derived. The AI model used these features from each 20-second interval to generate an ischemia probability. For each novice, a hybrid model learned a weighting between the novice’s and the AI model’s outputs, producing a weighted ischemia probability. Dataset-1 analyses used 10-fold patient-level cross-validation; dataset-2 analyses used 5-fold patient-level cross-validation. Performance was assessed using the area under the receiver operating characteristic curve (AUROC), the area under the precision-recall curve (AUPRC), the false-positive rate (FPR), precision, and calibration.

### Hybrid novice–AI system

The hybrid novice–AI system comprises two components: an AI model and a hybrid model (Figure 1). The AI model was an Extra Trees model trained using the 111 cEEG features extracted from each 20-second interval. It generated an ischemia probability for every interval and served as the AI model prediction. The hybrid model was a random forest model trained separately for each novice monitor. Unlike the AI model, the hybrid model did not directly predict ischemia. Instead, it learned when the novice monitor was likely to be more reliable than the AI model. For each interval, the hybrid model used three inputs: the AI model ischemia probability, the AI model uncertainty, and a cEEG variability score. For an AI model probability *p_AI_*, uncertainty was quantified using Shannon entropy: *H*(*p_AI_*) *= -p_AI_ log*(*p_AI_*) *-* (*1 - p_AI_*) *log*(*1 - p_AI_*). The cEEG variability score summarized how heterogeneous the 111-dimensional feature vector was within an interval. For interval *i*, with feature vector ***x_i_*** ∈ ***R***^111^, the variability score was computed as: *v_i_ = 0.5 × std(x_i_*) *+ 0.5 × range*(*x_i_),* where *std*(.) and *range*(.) denote min–max normalization using statistics estimated from the training data.

**Figure 1.**
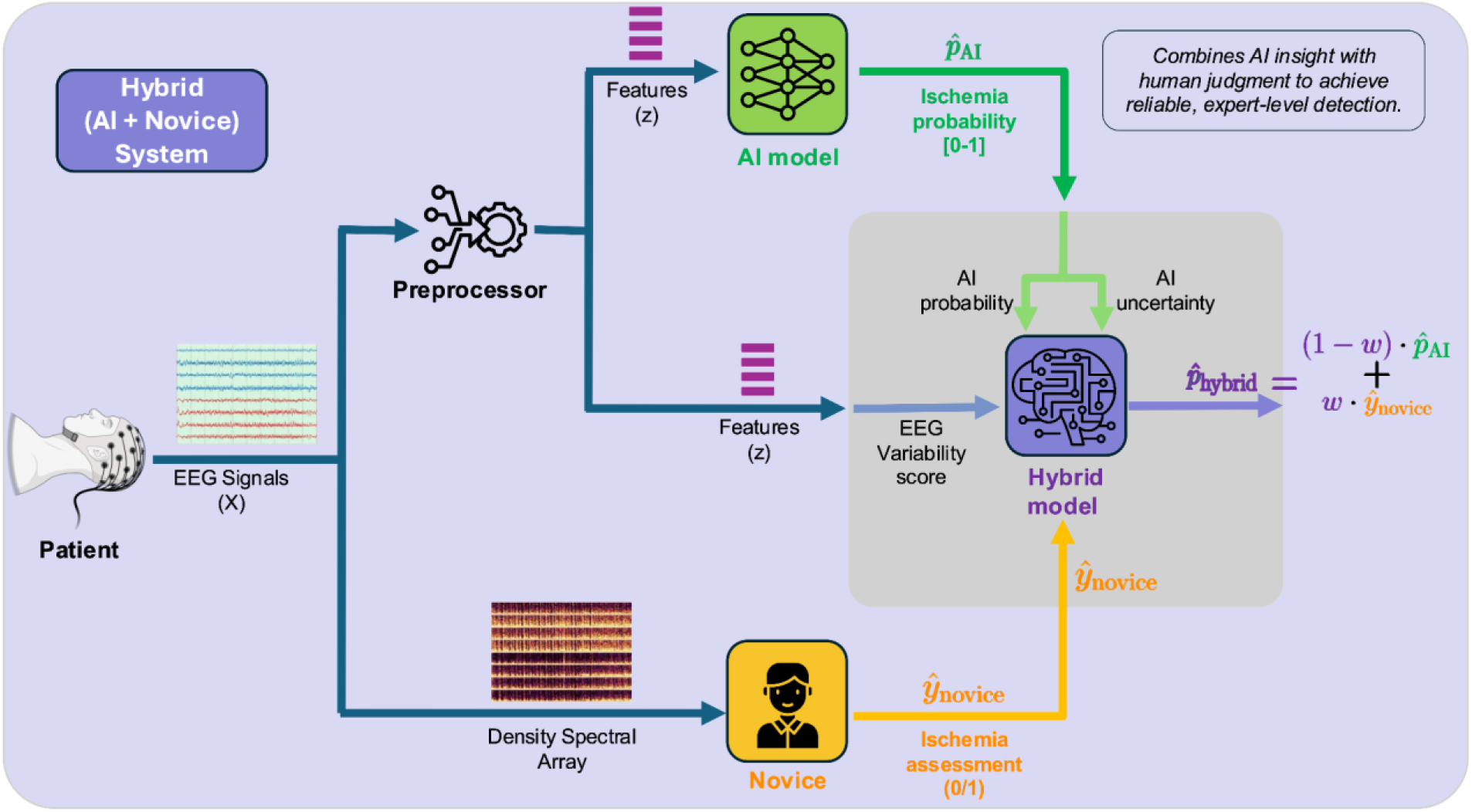
Schematic of the components of the hybrid novice-AI system. cEEG signals are presented to the AI model, the hybrid system, and a novice. A preprocessor extracts features from 20-second intervals of cEEG signals, which are provided as inputs to both the AI model and the hybrid system. Additionally, the density spectral array (DSA), derived from the cEEG signals, is provided to the novice. The DSA is a time-frequency visualization of cEEG signals that shows how spectral power changes over time, enabling rapid identification of ischemia (see Figure 2). The AI model outputs an ischemia probability *p*_AI_, which is decomposed into an AI probability and a corresponding uncertainty measure for use by the hybrid system. In parallel, a cEEG variability score is derived from the cEEG signals, and the novice ischemia assessment *y*_novice_ is passed directly to the hybrid system. The hybrid system employs a random forest model that integrates these four inputs—AI probability, AI uncertainty, cEEG complexity, and novice assessment—to produce a final probability of ischemia *p*_hybrid_.

The hybrid model was trained to predict whether the novice monitor was more accurate than the AI model for a given interval. The target label was positive when the novice assessment was correct, and the thresholded AI model prediction was incorrect; otherwise, it was negative. This allowed the hybrid model to learn interval-level patterns of disagreement between the novice monitor and the AI model. The hybrid model generated a novice-reliance weight *w* ∈ [0,1], where larger values indicated greater reliance on the novice monitor and smaller values indicated greater reliance on the AI model. The final hybrid probability was computed as: *p_hybrid_ = w×y_novice_ +* (*1-w*)*×p_AI,_* where *y_novice_* is the novice monitor’s binary assessment, and *p_AI_* is the AI model probability. The AI model was treated as fixed after training; the hybrid novice–AI system modified only how the AI model output and novice assessment were combined. The full target-label construction is shown in Table S5 in the Supplementary Information. Robustness across alternative AI models is reported in Section S9 in the Supplementary Information.

### AI and novice performance

On dataset-1, at the F1-optimal threshold, the AI model achieved a sensitivity of 0.60, a precision of 0.43, and a specificity of 0.98. In contrast, novices demonstrated higher sensitivity and precision, with a slight increase in FPR and marked heterogeneity across individuals. For example, novice sensitivities ranged from 0.54 to 0.83. This systematic disagreement, where the AI and novices make different types of errors, establishes a complementary error structure that the hybrid novice–AI system can leverage to improve performance.

### Representative example of hybrid predictions

We first present a representative example illustrating the behavior of the novice, AI, and hybrid system, followed by a comparison of their performance. Figure 2 shows a representative intraoperative cEEG segment with ischemic periods defined by expert consensus. The novice is highly sensitive to subtle ischemic changes, leading to false-positive predictions, whereas the AI model tends to miss mild ischemic changes, resulting in false-negative assessments. The hybrid systems adaptively integrate outputs from the AI model and the novice to produce predictions that more closely align with the expert determination. Additional illustrative cases demonstrating robustness to common cEEG artifacts are provided in Section S7 in the Supplementary Information.

**Figure 2.**
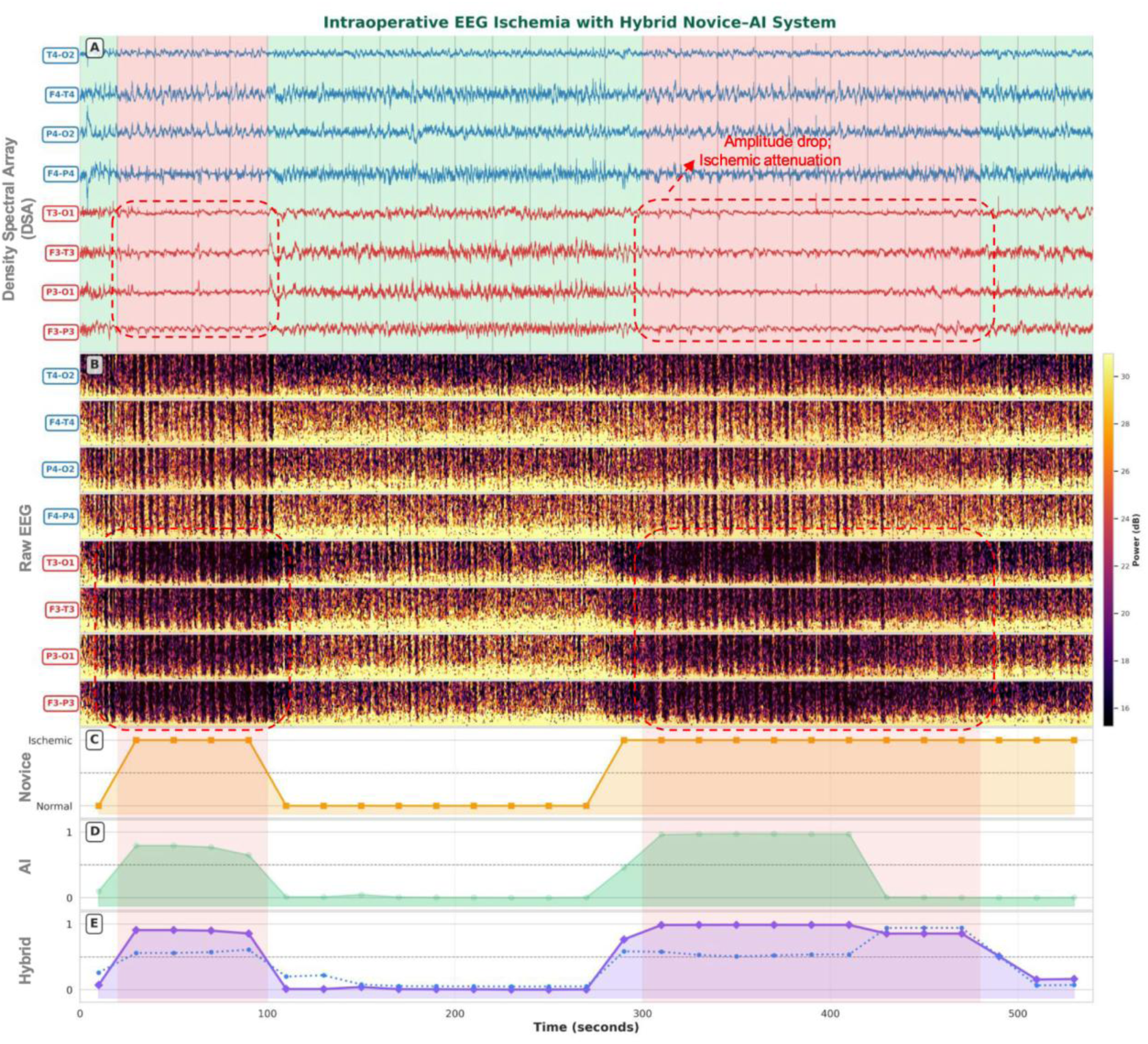
Balancing over-alerting novices and under-calling AI: hybrid systems align closely with expert cEEG interpretation. A. Plot of cEEG signals over a period of 500 seconds from eight channels, with four channels from the left hemisphere (blue labels) and from the right hemisphere (red labels). Pink-colored regions denote time periods labeled as ischemic by the expert neurophysiologist. Red dashed boxes highlight decreases in cEEG signal amplitude indicative of ischemic changes in the right hemisphere. B. Plot of density spectral array (DSA) derived from the cEEG signals in panel A. A DSA is a time–frequency visualization derived from cEEG signals that displays how spectral power evolves over time, enabling rapid identification of ischemia and is available to both experts and novices. Red dashed boxes highlight ischemic changes and are more clearly visible on DSA than on cEEG signals. C. Assessment by a novice who has identified the periods of ischemia and the healthy state based on the cEEG signals and the DSA. D. Predictions of the probability of ischemia by the AI model, generating a prediction every 20 seconds. E. Predictions of the probability of ischemia (shown in purple) by the hybrid system that dynamically combines the novice assessment with the AI model probability, generating a prediction every 20 seconds. The dotted blue line indicates the weight assigned by the hybrid system to the novice.

### Non-inferiority to experts

Using dataset-2, we evaluated whether the hybrid systems operated within the range of natural inter-expert variability in intraoperative cEEG interpretation. Figure 3 presents results from a non-inferiority analysis comparing hybrid systems (purple) and novice-only performance (yellow) with expert performance. Panel A shows non-inferiority bounds for delta sensitivity and delta FPR derived from expert variability. Panel B shows that the performance of hybrid systems falls within the bounds, indicating that they are statistically noninferior to expert-level performance. In contrast, novice-only performances fall outside these bounds, indicating that novices alone cannot achieve expert-level performance.

**Figure 3.**
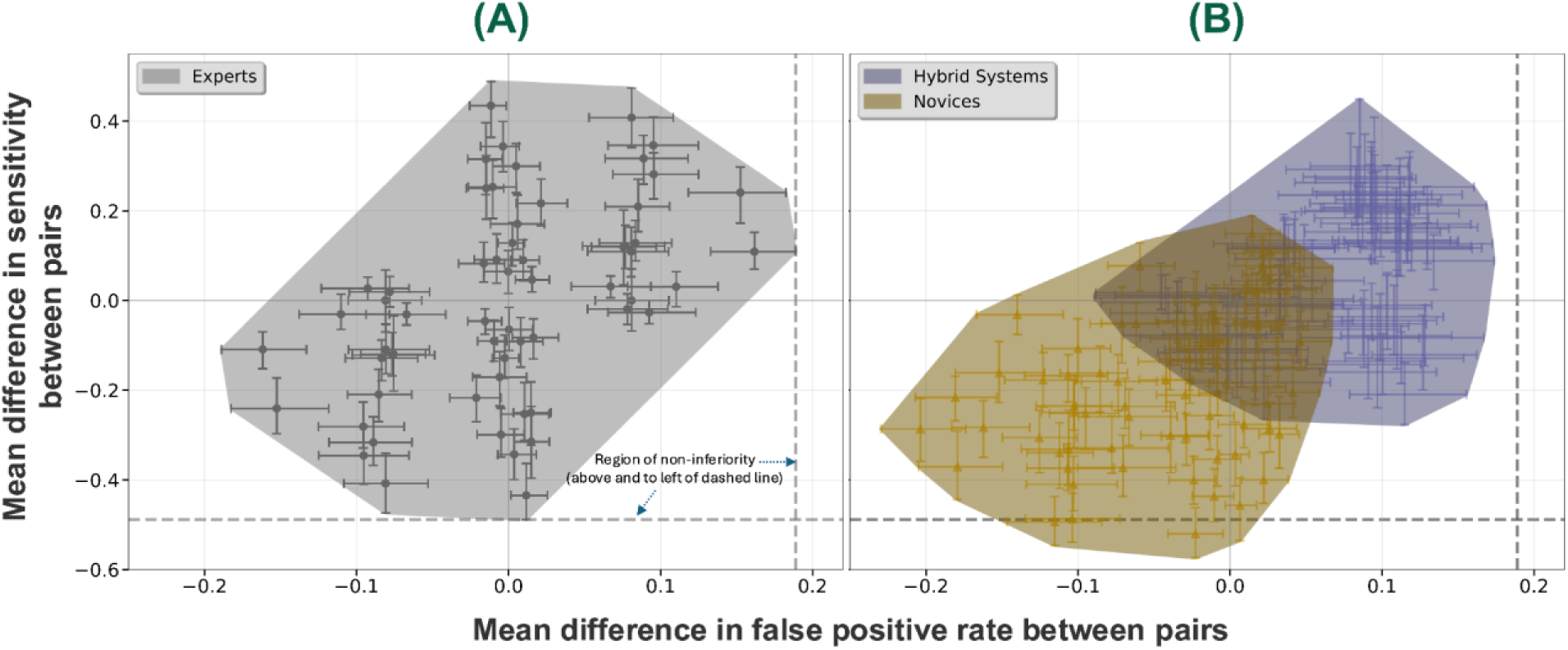
Plots of pairwise differences of sensitivity and false positive rate for expert neurophysiologist pairings, novice-AI hybrid system pairings, and novice pairings. A. Plot of the pairwise differences of sensitivity and false positive rate for expert neurophysiologist pairings to demonstrate inter-expert differences. Each point represents the mean difference in sensitivity (x-axis) and false-positive rate (y-axis) between a pair of experts, with error bars indicating 95% bootstrap confidence intervals. The horizontal and vertical dashed lines represent the non-inferiority bounds for sensitivity and false-positive rate, respectively. B. Plot of the pairwise differences of sensitivity and false positive rate for hybrid system pairings (in blue) and novice pairings (in orange). Each blue (orange) point represents the mean difference in sensitivity (x-axis) and false-positive rate (y-axis) between a pair of hybrid systems (novices), with error bars indicating 95% bootstrap confidence intervals. The horizontal and vertical dashed lines are the same as in A. The hybrid systems lie fully within the non-inferiority region, whereas the novices cross the horizontal bound.

### FPR at high sensitivity

Figure 4 compares the ROC curves and FPR performance of the hybrid systems with those of the AI model and the novices. In Panel A, the ROC curves for all hybrid systems outperform those of the AI model, demonstrating superior ROC performance across the clinically relevant operating range (sensitivity ≥ 0.80). Consistent with this, the hybrid systems achieved AUROC values of 0.967–0.971 compared with 0.957 for the AI model. Panel B shows that, at four fixed sensitivity thresholds (0.80, 0.85, 0.90, and 0.95), the hybrid systems consistently achieve lower FPRs than the AI model. At 80% sensitivity, the hybrid systems reduced the mean FPR by approximately half compared with the AI model (≈0.02 vs ≈0.04), with a smaller but still meaningful advantage retained at 90% sensitivity (≈0.07 vs ≈0.095). Two novices failed to achieve a sensitivity ≥ 0.80, and none achieved > 0.90. Collectively, these findings indicate that hybrid systems can simultaneously achieve clinically adequate sensitivities and acceptable FPRs.

**Figure 4.**
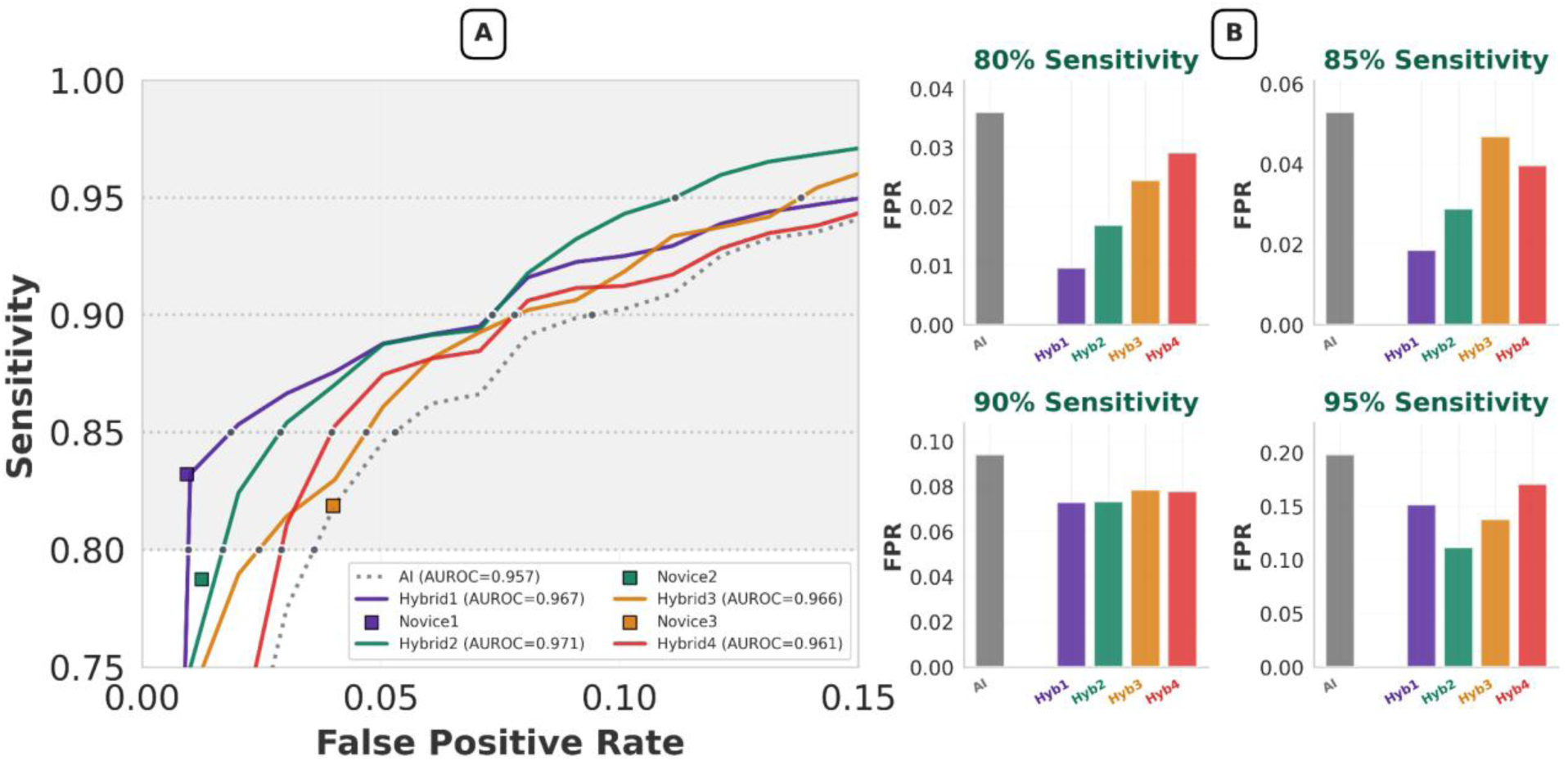
Performance on the receiver-operating characteristic (ROC) curves for the four hybrid systems compared to the AI system. A. Mean receiver-operating characteristic (ROC) curves for the AI system and the four hybrid systems are presented for the novices labeled N1, N2, N3, and N4. The ROC curves of all hybrid systems dominate those of the AI system. The three squares on the plot represent the sensitivity and false positive rate (FPR) for novices N1, N2, and N3. Novice N4 is not displayed on the plot because its sensitivity is 0.54, which falls below 0.75. The horizontal dashed lines denote sensitivities of 80%, 85%, 90%, and 95%. B. Bar plots of the FPRs of the AI system (grey color) and the four hybrid systems at varying sensitivities of 80%, 85%, 90%, and 95%. All hybrid systems achieve lower FPRs than the AI system across the four sensitivities.

### Precision at high sensitivity

A similar pattern was observed in the precision–recall domain (Figure 5). The PR curves for all the hybrid systems were well above those of the AI model, indicating higher precision across the full recall range. Improvements over the AI model were particularly pronounced: while the AI model achieved an AUPRC of 0.546, the four hybrid systems achieved AUPRC values of 0.726, 0.676, 0.647, and 0.610. These gains were substantially larger than those observed in ROC space, a divergence expected in rare-event settings such as ours (ischemia prevalence ≈3%). Although some novices achieved comparable precision at specific thresholds, these outcomes were achieved at much lower sensitivities, thereby limiting clinical utility. In the clinically relevant high-sensitivity region, the hybrid systems consistently outperformed both novices and the AI model, delivering more reliable detection, fewer false alarms, and more true-positive identifications.

**Figure 5.**
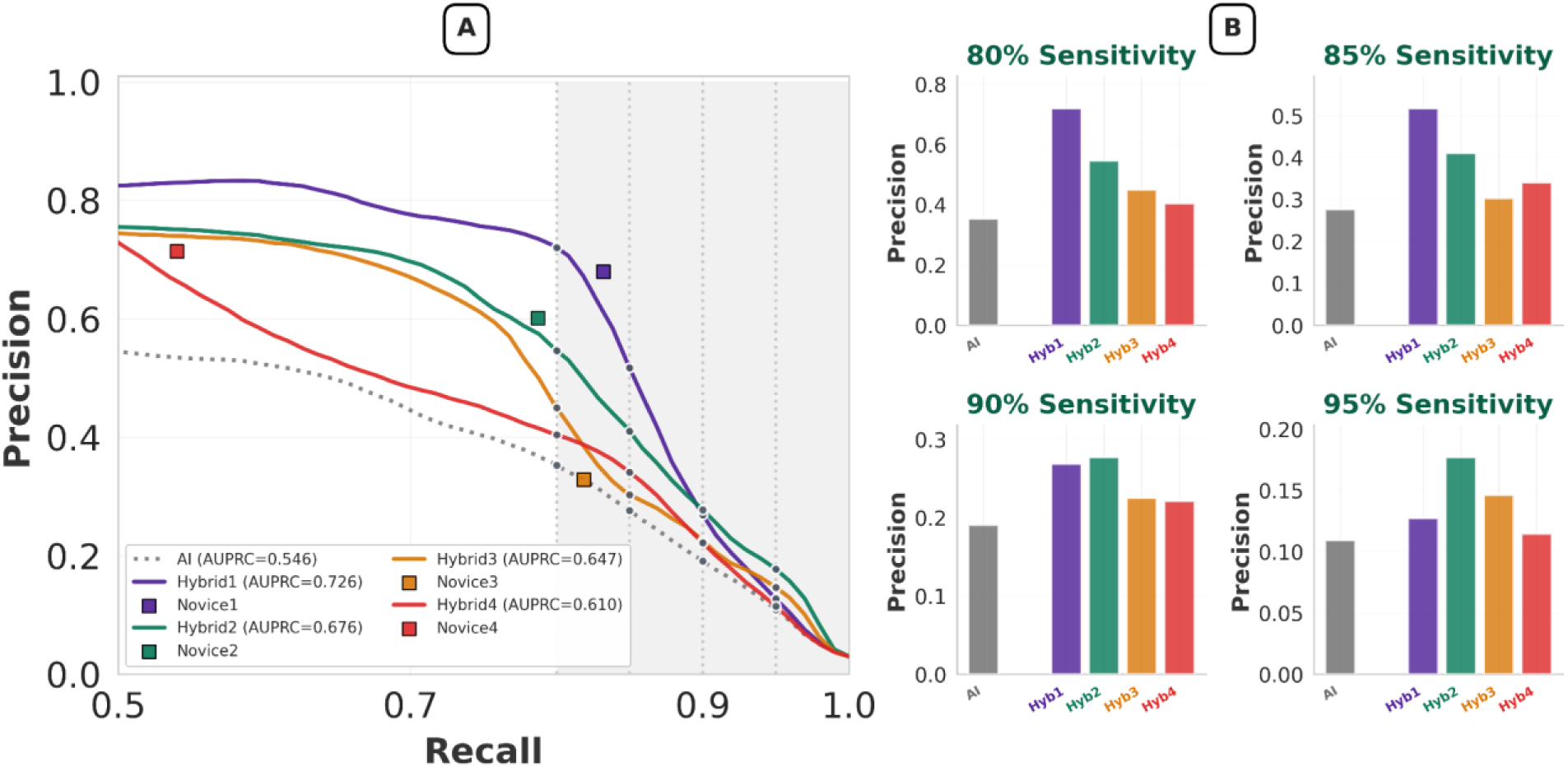
Performance on the precision-recall (PR) curves for the four hybrid systems compared to the AI system. A. Mean precision-recall (PR) curves for the AI system and the four hybrid systems are presented for the novices labeled N1, N2, N3, and N4. The PR curves of all the hybrid systems dominate those of the AI system. The four squares on the plot represent the precision and recall for novices N1, N2, N3, and N4. The vertical dashed lines denote sensitivities of 80%, 85%, 90%, and 95%. B. Bar plots of the precisions of the AI system (grey color) and the four hybrid systems at sensitivities of 80%, 85%, 90%, and 95%. All hybrid systems achieve higher precision than the AI system across the four sensitivities.

### Calibration

We evaluated the calibration of the AI and hybrid systems’ probabilistic outputs on dataset-1 using the Brier score and log loss (Table 1). The hybrid system consistently achieved a lower mean Brier score and log-loss than the AI model, indicating that integrating novice input does not degrade probabilistic calibration and may yield modest improvements in probability accuracy. Additional bin-based calibration metrics are reported in Section S8 in Supplementary Information. These results show that the hybrid approach maintains calibration while yielding modest improvements in probabilistic accuracy.

**Table 1.** Calibration performance of AI and hybrid systems. Values are reported as mean ± standard deviation based on 200 paired bootstrap replicates. For each replicate, we calculated the differences in calibration performance between the hybrid and AI systems using the same resampled data. We assessed statistical significance using a paired bootstrap procedure, in which p-values were computed as the proportion of bootstrap replicates in which the AI system outperformed the hybrid system. Significance was assessed at p < 0.025, indicating that the hybrid system consistently outperformed in at least 97.5% of bootstrap replicates. Lower values signify better calibration for all reported metrics.

| Metric | AI (mean ± SD) | Hybrid (mean ± SD) |
| --- | --- | --- |
| Brier score | 0.016 ± 0.001 | 0.013 ± 0.002 (p<0.001) |
| Log loss | 0.057 ± 0.003 | 0.049 ± 0.006 (p<0.025) |

## DISCUSSION

Pairing an AI model with novices, individuals with minimal neurophysiology training, achieved expert-level performance for detecting intraoperative ischemia during CEA. The hybrid novice-AI system was statistically noninferior to experts, satisfying a key safety criterion for clinical translation.

The hybrid approach reduced FPR relative to the AI model, which is essential for real-world deployment, as frequent false alarms can distract the surgical team and erode trust in the monitoring system (Figures 4–5). Importantly, this reduction in FPR was achieved while maintaining clinically relevant sensitivity (≥0.80), with the benefit still present at higher sensitivities (e.g., 0.90). We also observed a clear advantage in PR space (Figure 5), reflecting better precision at a given sensitivity. This is particularly important in rare-event settings such as ours (≈3% ischemia prevalence), where AUROC can remain uniformly high despite meaningful differences in minority-class performance, whereas AUPRC more directly reflects improvements in precision. Together, these findings demonstrate that the hybrid system can close the performance gap between novices and experts while simultaneously reducing unnecessary alerts compared with AI alone, supporting safer and more practical IONM.

The clinical utility of an IONM system relies on the reliability of its probabilistic outputs. Well-calibrated probabilities are essential for decision-making and risk communication in the operating room. The hybrid systems showed improved probability calibration compared with the AI model, as reflected by lower Brier scores and log-loss values. These calibration gains were accompanied by improved discrimination, suggesting that integrating novice input reduces overconfidence in the AI model’s predictions. The AI model captures subtle signal patterns, while novices help reduce false positives, resulting in more accurate and clinically reliable predictions. We also observe more stable behavior in the presence of common intraoperative artifacts, as shown in Section S7 in the Supplementary Information.

Importantly, despite variability in novice performance (N1–N4), the hybrid system consistently outperformed the AI model. As expected, combining the AI model with a stronger novice yielded greater overall gains in sensitivity, specificity, and precision (see Figures 4-5). Further analyses showing how the hybrid system learns when to rely more on the novice than on the AI are provided in Section S6 in the Supplementary Information.

Our findings suggest four key implications. First, a hybrid novice–AI system can enhance IONM by enabling novices to achieve expert-level performance, reducing the need for expert oversight. Second, lower FPRs, combined with high sensitivity, may alleviate alarm fatigue and cognitive burden while ensuring timely detection of ischemia, which is crucial for intraoperative decisions. Third, improved calibration enables the use of hybrid systems in workflows that require reliable confidence estimates. Fourth, hybrid systems enable structured task-sharing in which a novice monitors the patient with AI support, thereby improving overall performance and reducing the need for expert monitoring during surgery. A mechanistic analysis of why the hybrid system performs reliably is provided in Section S10 in the Supplementary Information.

This study draws data from multiple hospitals within a large academic health system, capturing substantial real-world heterogeneity in patients, workflows, and intraoperative practice. Additional validation in different health systems is essential for broader applicability across varied patient populations and practices. While external validation remains important, prospective real-world deployment will provide the ultimate test of clinical utility. Our study establishes a proof of concept for hybrid monitoring and lays the groundwork for broader studies examining workflow integration, resource requirements, and operational feasibility across diverse surgical settings (16,17).

This study has several limitations. Non-inferiority analyses relied on 30 expert-annotated patients; although bootstrap estimation mitigated sample constraints, larger expert datasets will better define the precision of estimates. Novice annotations were performed in a laboratory setting rather than in the operating room. Downstream outcomes such as time-to-detection or shunt placement decisions were not assessed and warrant prospective evaluation.

## Conclusion

Hybrid novice–AI systems achieved expert-level ischemia detection while delivering lower FPR and higher precision at clinically relevant sensitivities than either the AI model or the novices alone. Beyond gains in discrimination, the hybrid system maintained reliable probabilistic outputs and showed evidence of modest improvements in calibration. This combination of accurate detection and trustworthy confidence estimates is particularly critical for intraoperative decision-making, where alerts must be timely and dependable. Collectively, these findings delineate a pathway for extending reliable intraoperative cEEG monitoring in environments with limited expert neurophysiological coverage. This is a proof-of-concept study, and next steps include multi-site validation, careful workflow integration, and prospective evaluation in the operating room.

## METHODS

### Datasets

We used two intraoperative cEEG datasets from patients undergoing CEA between 2009 and 2017. Dataset-1 included data from 400 patients, 28 of whom exhibited intraoperative ischemic changes, and was used for primary performance and calibration analyses. Dataset-2 comprised 30 patients, 15 of whom exhibited intraoperative ischemic changes, and was used for expert comparison and non-inferiority analysis. The cEEG data were collected across multiple hospitals within a large U.S. academic health system comprising more than 40 hospitals, capturing real-world variability in clinical practice.

The cEEG signals were acquired at 500 Hz using XLTEK systems (Natus Medical Inc.) from eight subdermal electrodes positioned according to the international 10–20 system. These provided four bipolar channels per hemisphere: F3–P3, P3–O1, F3–T3, and T3–O1 on the left; and F4–P4, P4–O2, F4–T4, and T4–O2 on the right. Voltage recordings were obtained in a bipolar montage, representing the potential difference between electrode pairs, without subsequent re-referencing. During signal acquisition, the analog front end included a hardware band-pass filter from 0.5 to 70 Hz and a 60 Hz notch filter to reduce line noise and slow drift. For preprocessing before modeling, signals were filtered using a digital fourth-order Butterworth band-pass filter with a passband from 0.5 to 45 Hz. Additional acquisition details are provided in Table S2 in the Supplementary Information.

From each recording, we extracted 10 minutes of cEEG signals following the clamping of the carotid artery, when the risk of ischemia is highest. Signals were partitioned into 20-second non-overlapping intervals, yielding 12,000 intervals in dataset-1 and 900 intervals in dataset-2. In dataset-1, periods of ischemia were determined based on annotations by the neurophysiologist who performed the intraoperative monitoring. In dataset-2, periods of ischemia were determined by five experts. Dataset-1 included 285 ischemic intervals (2.38%), whereas dataset-2 had a higher ischemic interval fraction of 17.78%. Model training and evaluation were performed using these 20-second intervals. Table 2 summarizes patient demographics and key dataset characteristics.

**Table 2.** Patient and dataset characteristics for the two intraoperative cEEG datasets.

| Characteristic | Dataset-1 | Dataset-2 |
| --- | --- | --- |
| **Data type** | Intraoperative cEEG | Intraoperative cEEG |
| **Patients (Intervals)** | 400 (12,000) | 30 (900) |
| **Age (years)** | Median (IQR): 70 (64-77)<br>Range: 23-96 | Median (IQR): 70 (64-77)<br>Range: 23-96 |
| **Sex** | Male: 206 (6180), 51.5%<br>Female: 194 (5820), 48.5% | Male: 16 (480), 53.3%<br>Female: 14 (420), 46.7% |
| **Side of surgery** | Right: 214 (6420), 53.5%<br>Left: 186 (5580), 46.5% | Right: 17 (510), 56.7%<br>Left: 13 (390), 43.3% |
| **Period** | January 2009 - December 2017 | January 2009 - December 2017 |
| **Label type** | Intraoperative labels, novice labels | Expert labels, novice labels |
| **Role in study** | Primary performance analysis (Figures 4–5: ROC, PR curves, sensitivity, FPR, precision, calibration) | Expert comparison and non-inferiority analysis (Figure 3) |

**Abbreviations:** cEEG = continuous electroencephalography; IQR = interquartile range

### Label definition

Labels for the 20-second intervals were obtained in two ways. In dataset-1, periods of ischemia were obtained from intraoperative notes maintained by the expert neurophysiologist performing IONM. For each 20-second interval, we derived a positive, or ischemic, label by superimposing the interval on the note-derived periods of ischemia. An interval was labeled positive if more than half of its duration, defined as at least 10 seconds, coincided with a period of ischemia. In dataset-2, five expert neurophysiologists independently reviewed the cEEG signals while blinded to intraoperative notes, yielding five expert labels per interval. A consensus label was defined by majority vote among the experts. Four novice monitors also labeled both dataset-1 and dataset-2, as described below. Inter-rater agreement among experts and novices is reported in Section S5 in the Supplementary Information.

### Novice participants: selection, training, and performance calibration

We recruited four engineering and data science trainees with no formal background in cEEG, interpretation, and IONM. All novices received structured training focused on recognizing canonical cEEG patterns associated with cerebral ischemia, rather than on comprehensive cEEG interpretation. The training comprised two sessions: (1) an introductory session by an expert neurophysiologist that covered common ischemic cEEG patterns and common confounders; (2) a practice session in which a novice labeled a set of cEEG recordings with assistance and feedback from an expert. Each novice labeled ischemic intervals in both datasets using a custom forward-only cEEG viewer that advanced automatically every 20 seconds to simulate real-time IONM, with a limited look-back window (≤120 seconds). For each patient, novices labeled a 10-minute pre-clamp period followed by a 10-minute post-clamp period. Novices were not provided with expert labels or intraoperative notes.

### Feature construction

From each 20-second interval, we extracted quantitative cEEG features representing spectral, amplitude, and hemispheric symmetry characteristics. Power spectral density was estimated within each interval using Welch’s method with a 2-second Hamming window, 50% overlap, and 0.5-Hz frequency resolution. Power values were expressed in decibels to stabilize variance. We computed absolute band powers in five frequency bands: delta (0.5–4 Hz), theta (4–8 Hz), alpha (8–12 Hz), beta (12–30 Hz), and gamma (30–45 Hz). We also computed three band-power ratios: the alpha–delta ratio (ADR), the beta–delta ratio (BDR), and the alpha–beta–delta–theta ratio (ABDTR). These ratios were included to capture spectral slowing, which is a common cEEG change during cerebral ischemia. Additional features included spectral edge frequency at 90% power (SEF90), amplitude-integrated EEG (aEEG), and a pairwise-derived brain symmetry index (pdBSI). SEF90 was defined as the frequency below which 90% of the total spectral power was contained. aEEG summarized the amplitude range of the smoothed cEEG envelope within each interval. pdBSI measured interhemispheric asymmetry by comparing spectral power between homologous left- and right-sided channel pairs.

For each channel, we computed 10 spectral and amplitude features, yielding 80 channel-level features. We also computed 10 features that averaged the 10 channel-level features from the four left-hemisphere channels, 10 features that averaged the 10 channel-level features from the four right-hemisphere channels, and 10 features that averaged the 10 channel-level features from all eight channels, yielding 30 averaged features. Finally, a single pdBSI feature was computed, bringing the total to 111 features per interval. To account for patient-specific baseline differences, features were calculated as z-scores by normalizing raw feature values to a 120-second artifact-free pre-clamp baseline from the same patient. A complete list of features is provided in Tables S3–S4 in the Supplementary Information.

### Evaluation methods

We evaluated whether the AI and hybrid systems operated within the range of natural inter-expert variability in intraoperative cEEG interpretation. Using dataset-2, which includes multiple expert annotations, we evaluated the non-inferiority of the novice–AI systems relative to a panel of expert neurophysiologists. Following Scheuer and colleagues (18,19), we quantified inter-expert variability by treating each expert (R1–R5) in turn as the reference and computing pairwise differences in sensitivity and false-positive rate (FPR) relative to the remaining experts. This yielded empirical inter-expert difference distributions, from which non-inferiority bounds were derived. Specifically, we defined the non-inferiority region on the (Δsensitivity, ΔFPR) plane using (i) the lowest 95% confidence-interval whisker for sensitivity differences and (ii) the rightmost 95% confidence-interval whisker for FPR differences observed across expert–expert comparisons (18,19). A hybrid system was declared non-inferior if its entire 95% confidence interval lay within these bounds, indicating that its performance was no worse than the variability observed among experts. Figure 3 illustrates this criterion and summarizes the resulting non-inferiority analysis. Additional details are provided in Section S4 in the Supplementary Information.

To evaluate performance in clinically relevant operating regions, we focused on FPR and precision at fixed high sensitivities using dataset-1. Specifically, we evaluated the performance of the hybrid systems at sensitivity levels of 80–95%. For each novice, the corresponding hybrid system, and the AI, we computed the AUROC, the AUPRC, precision, and FPR at fixed sensitivities of 80%, 85%, 90%, and 95%. Given the low prevalence of ischemia, AUPRC may be particularly informative in this setting (see Section S9 in the Supplementary Information. We compared the performance of the hybrid systems with that of the novices and the AI when used individually. Figures 4 and 5 show the corresponding receiver operating characteristic (ROC) and precision–recall (PR) curves for the four novices, the four hybrid systems, and the AI model. We report both threshold-free metrics (AUROC and AUPRC) and threshold-based metrics (sensitivity, FPR, precision). For threshold-based metrics, we used nested cross-validation within the training data to select the F1-optimal threshold, which balances sensitivity and precision. The selected threshold was then applied unchanged to the held-out test data. To compare the AI and hybrid systems, we used a paired sign test. All evaluations were done using bootstrap resampling (200 replicates).

Calibration of probabilistic outputs was evaluated on dataset-1 by comparing the AI model’s predicted event probabilities with those of the corresponding hybrid novice–AI systems. For each system, we assessed agreement between predicted probabilities and observed event frequencies using the Brier score and log loss. The Brier score uses the mean squared error to assess the accuracy and calibration of probabilistic predictions. Log loss measures the negative log-likelihood of the true outcome under the predicted probabilities, penalizing confident but incorrect predictions and rewarding well-calibrated uncertainty. Lower values indicate better performance for both metrics.

### Experimental setup

We used two intraoperative cEEG datasets with distinct roles. For dataset-1, we obtained a single set of intraoperative ischemia labels. We used this dataset for the primary performance evaluation (Figures 4 and 5), including ROC and PR curves, sensitivity, FPR, precision, and calibration. We performed 10-fold patient-level cross-validation. For dataset-2, we obtained annotations from five expert neurophysiologists. We used this dataset for the non-inferiority analysis (Figure 3). We performed 5-fold patient-level cross-validation. To prevent information leakage during hyperparameter tuning and during adaptation of the hybrid novice–AI system to each novice, we used nested patient-level cross-validation. The AI model was trained only on training folds. Novices annotated held-out data that were not used to train the AI model. The hybrid model was trained only on the adaptation subset. None of the data used for training either model or for novice adaptation was included in the final evaluation dataset. Additional details are provided in Section S3 in the Supplementary Information.

### Statistical testing

All performance estimates were computed from held-out predictions using patient-level data splits to prevent information leakage. For analyses using interval-level predictions, uncertainty was estimated using patient-level bootstrap resampling with 200 replicates, preserving clustering of intervals within patients. For non-inferiority analysis, the comparisons of interest were hybrid systems versus expert neurophysiologists and novice monitors versus expert neurophysiologists. Non-inferiority was assessed using bootstrap 95% confidence intervals for differences in sensitivity and FPR. A system was considered non-inferior only if its full 95% confidence interval lay within the expert-derived non-inferiority bounds. For paired comparisons between the AI model and hybrid systems, we used paired tests on matched held-out predictions from the same intervals. Calibration was compared using paired bootstrap differences in Brier score and log loss. Statistical significance was assessed at an alpha level of 0.05. Tests were two-sided unless otherwise specified. Exact P values are reported where available.

## Ethics approval

The Institutional Review Board at the University of Pittsburgh approved this study. The cEEG data were de-identified, and novices had access only to these de-identified data.

## Data availability

The cEEG dataset used in this study is not publicly available due to patient privacy considerations and restrictions imposed by the data use agreement with the hospital system.

## Code availability

The code used for the analyses will be made publicly available at: https://github.com/batmanlab/EEG-Hybrid-AI-Systems upon publication.

## Acknowledgements

N.M. was supported by a National Institutes of Health (NIH) award (U24 TR004111). J.W.A. was supported by an NIH training program award (T15 LM007059). The views expressed in this article are those of the authors and not necessarily those of the NIH.

## Author contributions

N.M. led the study’s conceptualization, created the figures, and wrote the initial draft of the manuscript. N.M., H.S., J.W.A., and Y.R. reviewed the cEEG records and provided novice-level annotations. PDT collected and verified the raw data and provided expert-level annotations. H.C.-C. and H.K.A. assisted with the formal analysis. K.B. and S.V. supervised the project, contributed to the methodology, and reviewed and edited the manuscript. All authors had full access to the data and shared final responsibility for the decision to submit for publication.

