## Supplementary information for "Hybrid novice–AI system achieves expert-level performance in intraoperative ischemia detection"

The supplementary information provides further details that support the main findings. We first provide additional details of the intraoperative cEEG datasets, data acquisition protocols, and feature construction. We then detail the AI model and the formulation and training of the hybrid novice–AI system. Next, we establish expert-level performance using a non-inferiority framework grounded in inter-expert variability and characterize inter-rater agreement among novices and experts. Building on this foundation, we provide mechanistic insight into the hybrid novice–AI system's behavior through decision maps and uncertainty explainability analyses, followed by qualitative robustness evaluations under common intraoperative cEEG artifacts. We conclude with calibration analyses, robustness to alternative AI models, and an analysis demonstrating that structured decision-level diversity between AI models and novice monitors underlies the gains obtained by the hybrid system.

### List of Figures

S1 **Distribution of the per-patient percentage of ischemic intervals in dataset-1 and dataset-2.** (**a**) In dataset-2, the average percentage of ischemic intervals was 17.8%. This dataset showed significant variability among patients, with some individuals experiencing prolonged ischemia, whereas others had normal recordings. (**b**) In dataset-1, the average percentage of ischemic intervals was 2.4%. Most patients in this dataset exhibited normal recordings, which are typical in IONM practice. . . . . . . . . 9

S2 **Hybrid system decision maps for novice N1 (sensitivity-leaning, balanced behavior).** Heatmap color encodes the learned novice-reliance weight $w$, with warmer colors indicating greater reliance on the novice and cooler colors indicating greater reliance on the AI. Color transparency reflects data density, with darker regions indicating higher concentration of observed intervals (predominantly low cEEG complexity and low-to-moderate AI model uncertainty), and progressively lighter regions indicating sparser support toward higher complexity and uncertainty. The dominant structure is a strong gradient along the AI model uncertainty axis, with deferral increasing as the AI model becomes less confident. A secondary, but consistent, modulation is observed along cEEG complexity, particularly in the AI-uncertain regime, indicating increased reliance on N1 in harder conditions where complex signals coincide with AI model uncertainty and N1's sensitivity-leaning profile is most beneficial. 22

S3 **Hybrid system decision maps for novice N4 (conservative, precision-preserving behavior).** Heatmap color encodes the learned novice-reliance weight $w$, while color transparency reflects data density, highlighting that most observations lie in the low-complexity, low-to-moderate uncertainty region and that the upper-right portion of the map is sparsely populated. Deferral exhibits a strong monotone dependence on AI model uncertainty, with increasing reliance on N4 as the AI model becomes uncertain. In contrast to N1, modulation by cEEG complexity is comparatively weak, consistent with N4's conservative operating profile (high PPV and specificity, lower sensitivity) and the limited evidence for true positive events in high-complexity regimes. Accordingly, the hybrid system primarily leverages N4 to suppress false alarms under AI model uncertainty rather than escalating deferral solely based on signal complexity. . . . . . . . . . . . . . . . . . . . . . . . . . . . . . . . . . . . . . . . . . . 23

S4 **SHAP feature importance for AI predictive uncertainty.** Mean absolute SHAP values from an entropy-based uncertainty model using 16 aggregated cEEG features (4 frequency bands $\times$ 4 montages). Features are ordered by frequency band from low to high. Alpha- and theta-band features dominate uncertainty contributions, indicating that intermediate-frequency cEEG activity is the primary driver of AI predictive ambiguity. . . . . . . . . . . . . . . . . . . . 26

S5 **Electrocautery artifact.** An example illustrating the robustness of the hybrid system to electrocautery-induced artifacts. Repeated electrocautery generates high-amplitude noise in the right-hemispheric channels F4–T4 and F4–P4, which is evident in both the raw cEEG and the density spectral array (DSA). . . . . . . . . . . 28
S6 **Electrode pop artifact.** An example illustrating the robustness of the hybrid system to electrode pop artifacts. Dislodgement of the T3 electrode results in high-amplitude artifacts in the left-hemispheric channels F3–T3 and T3–O1, visible as high-frequency signals in the raw cEEG and uniformly elevated broadband power in the DSA. . . . . . 29
S7 **Anesthetic-induced artificat.** An example illustrating the robustness of the hybrid system in anesthetic-induced cEEG slowing. The anesthetic propofol produces diffuse cEEG slowing and reduced spectral power in the DSA across all channels, resembling ischemia. . . . . . . . . . . . . . . . . . . . . . . . . . . . . . . . . . . . . . . . . . . . . . 30
S8 **Benign interhemispheric asymmetry.** An example illustrating the robustness of the hybrid system in the presence of benign interhemispheric asymmetry. Mild, stable asymmetry in amplitude and spectral power across all channels. . . . . . . . . . . . . . . . . . . . . . . . . . . . . 31
S9 **Limitation.** An example illustrating the limitation of the hybrid system. When both the AI model and the novice are incorrect, the hybrid system combines their errors, yielding a false positive. . . . . . . . . 32
S10 **Decision-level similarity among AI models.** Pairwise Cohen's $\kappa$ computed between thresholded binary predictions from each AI models, ordered by architectural similarity and decision coherence. Higher values indicate more similar alarm decisions beyond chance. Clear block structure emerges: bagged tree ensembles and gradient-boosted trees exhibit high within-group agreement, whereas tabular deep learning models (FTT, TabNet) and especially MLP show progressively lower agreement, reflecting distinct inductive biases and decision patterns. . . . . . . . . . . . . . . . . . . . . . . . . . . . . 37

S12 **Hybrid novice-AI systems consistently outperform both the AI model and the novices across diverse AI models.** Across all evaluated AI models, each corresponding hybrid system—built on top of that same AI model—yields large and consistent improvements in discrimination, increasing AUPRC by approximately 15–40% relative to its underlying AI model. Despite substantial heterogeneity in AI architectures and performance, every hybrid system satisfies the expert non-inferiority criterion, whereas none of the novices do. These results show that the hybrid system consistently improves the specific AI model it is paired with, achieving expert-level behavior while substantially improving precision–recall performance in low-prevalence settings. Reported values are mean ± SD AUPRC. AUPRC denotes the absolute improvement of the hybrid over its underlying AI model. . . . . . . . . . . . . 35
S13 **Decision-level agreement between AI models and novices.** Average Cohen's $\kappa$ values summarizing agreement (left) across all novices for each AI model, and (right) across all AI models for each novice. . . . . . . . . . . . . . . . . . . . . . . . . . . . . . . . . . . . . . . . . . . . . . . . 38

#### List of Acronyms

| | |
|---|---|
| aEEG | Amplitude-integrated EEG |
| AI | Artificial Intelligence |
| ABDTR | (Alpha+Beta)/(Delta+Theta) Ratio |
| ADR | Alpha-to-Delta Ratio |
| AUPRC | Area Under the Precision–Recall Curve |
| AUROC | Area Under the Receiver Operating Characteristic Curve |
| BDR | Beta-to-Delta Ratio |
| CEA | Carotid Endarterectomy |
| CI | Confidence Interval |
| cEEG | Continuous electroencephalography |
| CSV | Comma-Separated Values |
| DSA | Density Spectral Array |
| ECE | Expected Calibration Error |
| FN | False Negative |
| FP | False Positive |
| FTT | FeatureTokenizer-Transformer |
| FPR | False Positive Rate |
| GradBoost | Gradient Boosting Machine |
| Hz | Hertz |
| LGBM | Light Gradient Boosting Machine |
| Log loss | Logarithmic loss (cross-entropy) |
| MCE | Maximum Calibration Error |
| MLP | Multilayer Perceptron |
| pdBSI | Pairwise-derived Brain Symmetry Index |
| PPV | Positive Predictive Value |
| PSD | Power Spectral Density |
| qEEG | Quantitative EEG |
| SD | Standard Deviation |
| $SEF_{90}$ | Spectral Edge Frequency (90%) |
| SHAP | SHapley Additive exPlanations |
| TN | True Negative |
| TP | True Positive |
| XGB | Extreme Gradient Boosting |
| $\Delta$ | Absolute difference or change (e.g., $\Delta$AUPRC, $\Delta\kappa$) |

#### S1 cEEG Acquisition and Datasets

In this section, we describe the acquisition of cEEG signals and the heterogeneity in the occurrence of ischemia across datasets.

The continuous electroencephalography (cEEG) recordings were obtained using XLTEK clinical cEEG systems (Natus Medical Inc.) at a sampling frequency of 500 Hz. Eight subdermal needle electrodes were placed according to the international 10–20 system, utilizing four bipolar derivations per hemisphere (F3–P3,

P3–O1, F3–T3, T3–O1 on the left; F4–P4, P4–O2, F4–T4, T4–O2 on the right). This anterior-posterior, bilateral electrode configuration ensured symmetrical cortical coverage across both hemispheres, which is particularly well-suited to detecting lateralized ischemic changes typical of CEA. The voltage recordings were obtained in a bipolar montage, representing the potential difference between electrode pairs, without any subsequent re-referencing.

During signal acquisition, the analog front end included a hardware band-pass filter operating between 0.5 and 70 Hz and a 60 Hz notch filter to reduce line noise and slow drift. For preprocessing prior to modeling, we applied a digital fourth-order Butterworth band-pass filter (0.5-45 Hz) to further suppress high-frequency noise and very low-frequency baseline fluctuations.

Table S1: **Percentage of ischemia intervals per patient in dataset-2.** Each patient's 10-minute cEEG recording comprises 30 consecutive 20-second intervals.

| Patient num. | Number of positive intervals | Percentage of positive intervals |
|---|---|---|
| 1 | 6 | 20.0 |
| 2 | 0 | 0.0 |
| 3 | 3 | 10.0 |
| 4 | 6 | 20.0 |
| 5 | 27 | 90.0 |
| 6 | 0 | 0.0 |
| 7 | 22 | 73.3 |
| 8 | 27 | 90.0 |
| 9 | 0 | 0.0 |
| 10 | 26 | 86.7 |
| 11 | 0 | 0.0 |
| 12 | 0 | 0.0 |
| 13 | 0 | 0.0 |
| 14 | 0 | 0.0 |
| 15 | 0 | 0.0 |
| 16 | 0 | 0.0 |
| 17 | 0 | 0.0 |
| 18 | 0 | 0.0 |
| 19 | 5 | 16.7 |
| 20 | 0 | 0.0 |
| 21 | 0 | 0.0 |
| 22 | 0 | 0.0 |
| 23 | 4 | 13.3 |
| 24 | 0 | 0.0 |
| 25 | 0 | 0.0 |
| 26 | 0 | 0.0 |
| 27 | 7 | 23.3 |
| 28 | 0 | 0.0 |
| 29 | 27 | 90.0 |
| 30 | 0 | 0.0 |

Figure S1 illustrates the heterogeneity of ischemia occurrence in dataset-2. Although the mean positive fraction was 17.8%, most patients had no ischemic intervals, whereas a minority exhibited extensive ischemic activity, resulting in a strongly right-skewed distribution.

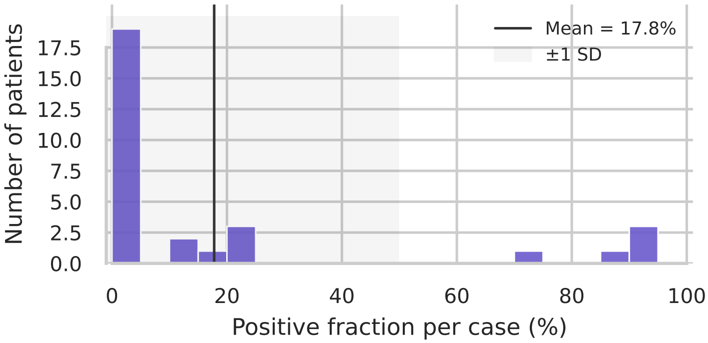

**(a)** Dataset-2 ($n$=30 patients)

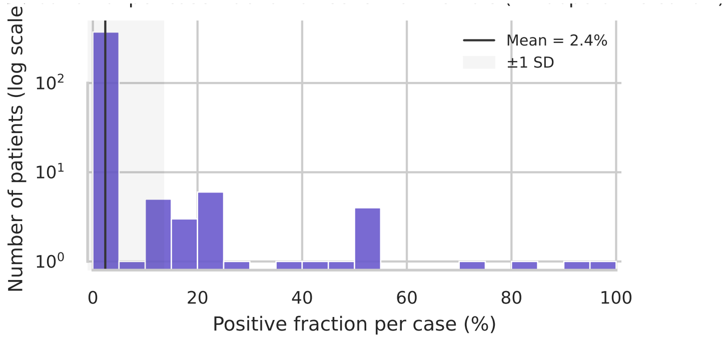

**(b)** Dataset-1 ($n$=400 patients)

Fig. S1: **Distribution of the per-patient percentage of ischemic intervals in dataset-1 and dataset-2.** (**a**) In dataset-2, the average percentage of ischemic intervals was 17.8%. This dataset showed significant variability among patients, with some individuals experiencing prolonged ischemia, whereas others had normal recordings. (**b**) In dataset-1, the average percentage of ischemic intervals was 2.4%. Most patients in this dataset exhibited normal recordings, which are typical in IONM practice.

Each patient's recording was exported as a binary cEEG file, accompanied by a time-synchronized text log containing annotations from the intraoperative neurophysiologist. The binary format was reverse-engineered to analyze the header structure, sample indexing, and signal encodings. The parsed data were then stored as comma-separated value (CSV) files for further analysis and aligned with the event logs to ensure label consistency.

In both dataset-1 and dataset-2 the cEEG signals were split into 30 consecutive 20-second intervals, totaling 10 minutes of recording per patient. All analyses were conducted at the interval level. In dataset-1, which included 400 patients, interval labels for ischemia were derived from the intraoperative expert neurophysiologist's annotations recorded during surgery. For dataset-2 of 30 patients, interval labels were defined by a majority-vote consensus among five expert neurophysiologists who independently reviewed the recordings retrospectively. In dataset-2, 17.78% of the intervals were ischemic. Table S1 summarizes the ischemic burden per patient, detailing the number and percentage of positive intervals for each patient. Figure S1 illustrates the variation in ischemia occurrence among patients in the dataset-2. While some patients did not exhibit any ischemic intervals, others experienced extensive ischemic activity.

Table S2: **Summary of cEEG acquisition parameters.**

| **Parameter** | **Description** |
|---|---|
| Acquisition system | XLTEK clinical cEEG (Natus Medical Inc.) intraoperative monitoring platform. |
| Sampling frequency | 500 Hz continuous recording. |
| Electrode type | Subdermal needle electrodes (disposable sterile stainless steel). |
| Electrode placement | Eight electrodes positioned according to the international 10–20 system.<br>Left: F3–P3, P3–O1, F3–T3, T3–O1;<br>Right: F4–P4, P4–O2, F4–T4, T4–O2. |
| Montage | Bipolar montage (anterior–posterior pairs per hemisphere); no re-referencing. |
| Analog filtering | Hardware band-pass 0.5–70 Hz; 60 Hz notch filter for power-line suppression. |
| Digital filtering | Fourth-order Butterworth band-pass (0.5–45 Hz) applied during preprocessing. |
| Data export format | Raw XLTEK binary files parsed to CSV with time-synchronized neurophysiologist annotations. |
| Interval segmentation | cEEG divided into non-overlapping 20-s intervals (30 per patient, ≈10 min total). |
| Purpose / downstream use | Provides standardized bilateral signals for 111 quantitative cEEG features (amplitude-, frequency-, and symmetry-based; see Section S2). |

### S2 cEEG Features

In this section, we describe the construction of 111 features from each 20-second interval, which are used by all AI and hybrid models.

##### Signal preprocessing

cEEG signals from each of the eight bipolar channels (F3–P3, P3–O1, F3–T3, T3–O1, F4–P4, P4–O2, F4–T4, T4–O2) were acquired at a sampling frequency of 500 Hz and band-pass filtered between 0.5 and 45 Hz using a fourth-order Butterworth design, consistent with the digital preprocessing described in Mina (12). Each 20-second interval therefore contained approximately 10,000 samples per channel. Signals were visually and automatically inspected for clipping, flat-lining, or high-variance artifacts (e.g., electrocautery noise). Intervals exhibiting gross artifacts were excluded from analysis. No rereferencing, resampling, or channel interpolation was performed.

#### Spectral estimation

Within each 20-s interval, the power spectral density (PSD) was estimated using Welch's method with a 2-s Hamming window (50% overlap) and 0.5-Hz frequency resolution, identical to the configuration validated in Mina's dissertation. Power values were expressed in decibels ($10 \times \log_{10}$ power) to stabilize variance and allow ratio computation. The PSD provided the basis for five absolute band-power measures, three spectral ratios, one spectral edge frequency, one amplitude-integrated measure, and one symmetry index, totaling ten base feature types.

#### Frequency bands

Band limits followed standard intraoperative cEEG conventions and the analog filter limits (0.5–70 Hz hardware; 0.5–45 Hz digital): $\delta$ (0.5–4 Hz), $\theta$ (4–8 Hz), $\alpha$ (8–12 Hz), $\beta$ (12–30 Hz), $\gamma$ (30–45 Hz). Power in each band, $P_b$, was computed as the integrated PSD across frequencies in that range:

$$P_b = \int_{f_1}^{f_2} \mathrm{PSD}(f)\, df.$$

These five band powers formed the foundation for all ratio and composite measures.

#### Band-power ratios

Three canonical ratios were included to quantify spectral slowing and loss of faster rhythms during ischemia, as established in prior CEA monitoring studies (12):

$$\mathrm{ADR} = \frac{P_\alpha}{P_\delta},$$

$$\mathrm{BDR} = \frac{P_\beta}{P_\delta},$$

$$\mathrm{ABDTR} = \frac{P_\alpha + P_\beta}{P_\delta + P_\theta}.$$

A reduction in these ratios reflects attenuation of higher-frequency activity relative to slow waves, a hallmark of hypoperfusion.

##### Spectral edge frequency

The spectral edge frequency (SEF) summarizes the upper bound of dominant energy. $\mathrm{SEF}_{90}$ was computed as the frequency $f^*$ satisfying

$$\int_{0.5}^{f^*} \mathrm{PSD}(f)\, df = 0.9 \int_{0.5}^{45} \mathrm{PSD}(f)\, df,$$

i.e., 90% of total spectral power lies below $f^*$. Decreases in $\mathrm{SEF}_{90}$ indicate spectral slowing and have been used intraoperatively as markers of hemispheric ischemia.

##### Amplitude-integrated EEG (aEEG)

Following the implementation in Mina (12), each 20-s segment was rectified, smoothed with a 2-s moving average, and displayed on a semi-logarithmic amplitude scale. The aEEG feature was defined as the difference between the upper and lower envelope values (max–min) over the 20-s interval, representing the dynamic amplitude range of background cEEG activity. This measure complements the spectral indices by capturing amplitude suppression during ischemia.

##### Pairwise-derived Brain Symmetry Index (pdBSI)

Hemispheric symmetry was quantified using the pairwise-derived Brain Symmetry Index (pdBSI) (12). For each homologous left–right pair ($i$), the normalized absolute difference in power spectra was computed:

$$\mathrm{pdBSI} = \frac{1}{N_f N_p} \sum_{i=1}^{N_p} \sum_{f} \frac{|P_{L,i}(f) - P_{R,i}(f)|}{P_{L,i}(f) + P_{R,i}(f)},$$

where $N_f$ is the number of frequency bins (0.5–45 Hz) and $N_p$ is the number of channel pairs. Higher pdBSI values indicate greater inter-hemispheric asymmetry, as typically observed during unilateral carotid cross-clamp ischemia.

##### Averaged features

The ten base features were computed for each of the eight bipolar channels (80 features). To summarize spatial patterns, identical computations were performed on averaged signals from the left hemisphere (F3–P3, P3–O1, F3–T3, T3–O1), the right hemisphere (F4–P4, P4–O2, F4–T4, T4–O2), and all eight channels combined (30 aggregate features). Finally, a single pdBSI feature was added, yielding 111 features per interval. This hierarchy preserved both localized and hemispheric information while maintaining a fixed-length input for machine-learning models.

Table S3: **Definitions of quantitative cEEG features with brief descriptions.**

| cEEG Feature | Symbol | Description |
|---|---|---|
| Delta power | $P_\delta$ | Integrated spectral power 0.5–4 Hz. |
| Theta power | $P_\theta$ | Integrated spectral power 4–8 Hz. |
| Alpha power | $P_\alpha$ | Integrated spectral power 8–12 Hz. |
| Beta power | $P_\beta$ | Integrated spectral power 12–30 Hz. |
| Gamma power | $P_\gamma$ | Integrated spectral power 30–45 Hz. |
| Alpha/Delta ratio | ADR | $P_\alpha / P_\delta$. |
| Beta/Delta ratio | BDR | $P_\beta / P_\delta$. |
| (Alpha+Beta)/(Delta+Theta) | ABDTR | $(P_\alpha + P_\beta)/(P_\delta + P_\theta)$. |
| Spectral edge frequency (90%) | $\mathrm{SEF}_{90}$ | Frequency below which 90% of total 0.5–45 Hz power resides. |
| Amplitude-integrated EEG | aEEG | Amplitude range (max–min) of smoothed cEEG envelope. |
| Pairwise-derived Brain Symmetry Index | pdBSI | Mean normalized absolute spectral difference between homologous left/right pairs. |

Table S4: **Summary of 111 cEEG features computed from each 20-second interval.**

| Type of feature | Number of features |
|---|---|
| 10 features derived from a single channel x 8 channels | 80 |
| 10 features derived from averaging 4 left-hemisphere channels | 10 |
| 10 features derived from averaging 4 right-hemisphere channels | 10 |
| 10 features derived from averaging all 8 channels | 10 |
| Pairwise-derived Brain Symmetry Index (pdBSI) | 1 |
| **Total** | **111** |

Together, these 111 features capture multiple physiologic dimensions of intraoperative cEEG dynamics: (1) absolute spectral power changes reflecting amplitude suppression; (2) band-ratio shifts indicative of slowing and desynchronization; (3) spectral edge shifts marking broadband frequency redistribution; (4) aEEG range representing overall voltage variability; and (5) pdBSI as a direct measure of hemispheric asymmetry.

#### S3 AI models and hybrid novice-AI system

The hybrid novice-AI system consists of two distinct AI models: the AI model and the hybrid model. Although we employed ExtraTrees for the AI model, we also evaluated other models (see Supplementary Section-S9).

#### AI Model

To develop the Extra Trees model, we utilized dataset-1 and applied a patient-level, stratified cross-validation strategy. The dataset was divided into 10 folds; in each iteration, 9 folds were used for training, and the remaining fold was reserved for testing. This iterative process ensured that each fold was used as a test set exactly once, yielding out-of-fold probabilistic predictions for every patient and interval in dataset-1. These predictions, derived from unseen data, were subsequently fed into the hybrid model.

Additionally, we trained a single Extra Trees model on dataset-1 and applied it to dataset-2 to generate probabilistic predictions for each patient and interval in the new dataset. This method mirrors a practical deployment scenario in which an AI model trained on a large historical cohort is used to assess new patients.

#### Hybrid Model

The hybrid model does not predict ischemia directly; instead, it assesses which source—the novice or the AI model—provides a more reliable prediction for each interval. The hybrid model uses the AI model's prediction and the 111 cEEG features as inputs, and it computes the following. First, the *AI model's probability* $\hat{p}_{\mathrm{AI}} \in [0, 1]$, which is the probability of ischemia output by the AI model. Second, the *AI model's uncertainty*, which is computed as the Shannon entropy of the AI model's probability. For a binary prediction with probability $\hat{p}_{\mathrm{AI}}$, uncertainty is defined as

$$H(\hat{p}_{\mathrm{AI}}) = -\hat{p}_{\mathrm{AI}} \log \hat{p}_{\mathrm{AI}} - (1 - \hat{p}_{\mathrm{AI}}) \log(1 - \hat{p}_{\mathrm{AI}}),$$

which is subsequently normalized by its maximum value $\log 2$ to yield a quantity in $[0, 1]$. Third, a *cEEG variability score*, which is a scalar summary of how heterogeneous or atypical the cEEG signals are in feature space. Let $\mathbf{x}_i \in \mathbb{R}^{111}$ denote the vector of 111 z-score–normalized cEEG features extracted from interval $i$. The cEEG variability score is computed as a normalized combination of the within-interval standard deviation and peak-to-peak range across feature dimensions:

$$v_i = \tfrac{1}{2}\,\widetilde{\mathrm{std}}(\mathbf{x}_i) + \tfrac{1}{2}\,\widetilde{\mathrm{range}}(\mathbf{x}_i),$$

where $\widetilde{\mathrm{std}}(\cdot)$ and $\widetilde{\mathrm{range}}(\cdot)$ denote min–max normalization using statistics estimated from the training data.

The target label for the hybrid model indicates when the novice performed correctly, while the AI model did not. As shown in Table S5, there are eight possible combinations of the AI model prediction, novice assessment, and ground truth; the last column indicates the target label for novice superiority over the AI model. This label helps the model identify instances in which the novice outperforms the AI.

The hybrid model is a Random Forest model trained with these target labels to generate a probabilistic output indicating the likelihood that the novice is superior to the AI model. This probability serves as a weighting factor, combining the novice's binary assessment with the AI model's probabilistic output to produce a final probability of ischemia for the interval, calculated as:

$$\text{Hybrid probability} = w \cdot \text{Novice} + (1 - w) \cdot \text{AI model}.$$

Throughout our experiments, the AI model was treated as fixed after training. Consequently, the hybrid novice-AI system focused solely on integrating the AI model's outputs with novice assessments, without modifying the AI model itself.

Table S5: **Construction of target labels for the hybrid model.** The binary label *Is novice superior to AI model?* indicates whether the novice outperformed the AI model on a cEEG interval.

| AI model prediction | Novice assessment | Ground truth | Is novice superior to AI model? |
|---|---|---|---|
| 0 | 0 | 0 | 0 |
| 0 | 0 | 1 | 0 |
| 0 | 1 | 0 | 0 |
| 0 | 1 | 1 | 1 |
| 1 | 0 | 0 | 1 |
| 1 | 0 | 1 | 0 |
| 1 | 1 | 0 | 0 |
| 1 | 1 | 1 | 0 |

#### S4 Non-Inferiority Evaluation

In this section, we discuss inter-expert variability and the non-inferiority evaluation we conducted to assess the performance of the novice-AI hybrid system compared to experts.

##### Inter-expert variability

We assessed inter-expert variability using the approach by Scheuer et al. (21; 22), involving five experts (R1–R5). Each expert served as the reference in turn, and we calculated pairwise differences in sensitivity and false-positive rate (FPR) for all other experts relative to the designated reference. This process produced an empirical inter-expert difference distribution that established the acceptable range for assessing non-inferiority.

**System-to-expert and novice-to-expert comparisons**

For each hybrid system and each novice, we calculated sensitivity and false-positive rate (FPR) relative to every expert used as a reference. For each predictor-expert pair, we determined the difference in sensitivity ($\Delta$sensitivity) and the difference in FPR ($\Delta$FPR) between the predictor and the reference expert. These paired differences were aggregated across 200 bootstrap replicates to obtain empirical mean differences and 95% confidence intervals (CIs). The CIs were defined by the 2.5th and 97.5th percentiles of the bootstrap distribution. Inter-expert variability was quantified similarly by computing expert-to-expert $\Delta$sensitivity and $\Delta$FPR values, with each expert serving as the reference in turn. This yielded empirical distributions of inter-expert differences that helped define the non-inferiority boundaries.

Predictors were declared non-inferior only if their entire 95% CI for $\Delta$sensitivity lay above the inter-expert sensitivity boundary and their entire 95% CI for $\Delta$FPR lay to the left of (i.e., did not exceed) the inter-expert FPR boundary. Aggregated results were visualized in two-dimensional non-inferiority plots in which each point represents the mean ($\Delta$sensitivity, $\Delta$FPR) with asymmetric 95% CI whiskers; convex-hull polygons summarize uncertainty envelopes for experts, novices, and hybrid systems, and dashed lines denote inter-expert non-inferiority boundaries. The region above (indicating higher sensitivity) and to the left (indicating lower FPR) of both boundaries is considered statistically non-inferior under this CI-based criterion. This method reflects Scheuer et al.'s "statistical Turing test," which treats performance indistinguishable from expert-to-expert variability as clinically equivalent to expert behavior.

#### S5 Inter-Rater Agreement

We quantified pairwise agreement among novices andexperts using Cohen's $\kappa$ (chance-corrected agreement) and raw percent agreement. Because ischemic intervals were rare in dataset-2 ($285/12000 = 2.38\%$ positives), we also report class-stratified agreement conditioned on the ground-truth label (agreement on true positives and true negatives). This avoids over-interpreting high overall agreement that can arise under severe class imbalance.

**Agreement among novices.**

Across the 12,000-interval novice-labeled cohort, novices exhibited moderate-to-substantial chance-corrected agreement (mean $\kappa = 0.61 \pm 0.11$ across the 6 novice pairs) despite very high raw agreement (mean overall agreement $97.34\% \pm 1.29\%$; Table S6). Importantly, agreement was markedly lower on true positives than true negatives (mean agreement on positives $76.37\% \pm 9.71\%$ vs. negatives $97.85\% \pm 1.41\%$; Table S8). This gap indicates that ischemic patterns are the primary source of ambiguity and inter-novice disagreement, whereas non-ischemic intervals are comparatively straightforward.

From the perspective of human-AI collaboration, the spread in novice $\kappa$ values (range $\approx$ 0.44–0.76 across pairs) supports the premise that novices are not interchangeable: they exhibit meaningfully different decision tendencies. This diversity is desirable for an adaptive hybrid system, because it demonstrates that performance gains are not merely due to learning a single stereotyped "novice pattern," but rather require robustness to heterogeneous novice behavior.

**Agreement among experts**

In dataset-2 (900 intervals), experts demonstrated moderate-to-substantial agreement (mean $\kappa = 0.67 \pm 0.12$ across 10 expert pairs; Table S7). Similar to novices, experts disagreed more on true positives than true negatives (positives $72.82\% \pm 15.92\%$ vs. negatives $92.61\% \pm 4.65\%$; Table S8), reinforcing that the positive class is inherently difficult even for trained monitors. The greater variability in expert agreement on the positive class indicates that borderline ischemic patterns may be interpreted differently by the experts, highlighting the challenges in interpreting subtle patterns.

**Novices vs. experts.**

Experts had a chance-corrected agreement than novices ($\Delta\kappa = +0.06$), whereas novices had higher overall and class-conditional raw agreement (Table S8). Under heavy class imbalance, raw agreement can be inflated by the dominant negative class and by conservative labeling strategies; therefore, we emphasize $\kappa$ for comparing consistency across groups, and we use class-stratified agreement to interpret where disagreements arise. Together, these results make two points relevant to our hybrid framing: (i) heterogeneity among novices is real and measurable (supporting the need for adaptive integration rather than a one-size-fits-all rule), and (ii) positive/ischemic intervals are the main locus of disagreement for *both* groups, motivating a hybrid strategy that can leverage complementary cues (e.g., AI confidence/uncertainty and novice judgments) to stabilize decisions on difficult positives. These results establish that both novices and experts disagree most strongly on positive/ischemic intervals and that novices exhibit heterogeneous operating tendencies.

#### S6 Qualitative Interpretation of Hybrid Decision-Making Behavior

In this section, we examine how differences between the novice and the AI help explain hybrid behavior. We show how the hybrid system adjusts how much it relies on the novice or the AI across different novices and levels of uncertainty. We focus on two complementary perspectives: (i) decision maps that visualize how the reliance of the hybrid system varies as a function of the uncertainty of the AI model and the complexity of cEEG between different novice operating

Table S6: **Pairwise agreement among novices on dataset-1** ($n$ = 12,000 **intervals;** 285 **positives,** 11,715 **negatives).**

| Pair | Cohen's $\kappa$ | Overall agreement | Agreement on positives | Agreement on negatives |
|---|---|---|---|---|
| N1 vs N4 | 0.71 | 98.68% | 70.53% | 99.37% |
| N1 vs N2 | 0.76 | 98.62% | 83.16% | 98.99% |
| N1 vs N3 | 0.55 | 96.18% | 86.67% | 96.41% |
| N4 vs N2 | 0.61 | 98.17% | 62.81% | 99.03% |
| N4 vs N3 | 0.44 | 95.88% | 70.53% | 96.50% |
| N2 vs N3 | 0.59 | 96.50% | 84.56% | 96.79% |

Table S7: **Pairwise agreement among experts on dataset-2** ($n$ = 900 **intervals;** 259 **positives,** 641 **negatives).**

| Pair | Cohen's $\kappa$ | Overall agreement | Agreement on positives | Agreement on negatives |
|---|---|---|---|---|
| R1 vs R2 | 0.69 | 90.11% | 76.45% | 95.63% |
| R1 vs R3 | 0.49 | 78.89% | 59.07% | 86.90% |
| R1 vs R4 | 0.62 | 85.44% | 60.62% | 95.48% |
| R1 vs R5 | 0.54 | 83.22% | 52.51% | 95.63% |
| R2 vs R3 | 0.59 | 83.00% | 68.73% | 88.77% |
| R2 vs R4 | 0.68 | 87.56% | 67.95% | 95.48% |
| R2 vs R5 | 0.64 | 86.44% | 61.39% | 96.57% |
| R3 vs R4 | 0.76 | 89.44% | 97.68% | 86.12% |
| R3 vs R5 | 0.74 | 88.78% | 91.89% | 87.52% |
| R4 vs R5 | 0.91 | 96.22% | 91.89% | 97.97% |

Table S8: **Summary of agreement (mean ± SD across all pairs) among novices (6 pairs) and among experts (10 pairs).**

| Metric | Novices | Experts |
|---|---|---|
| Cohen's $\kappa$ | $0.61 \pm 0.11$ | $0.67 \pm 0.12$ |
| Overall agreement | 97.34% ± 1.29% | 86.91% ± 4.75% |
| Agreement on positives | 76.37% ± 9.71% | 72.82% ± 15.92% |
| Agreement on negatives | 97.85% ± 1.41% | 92.61% ± 4.65% |

profiles, and (ii) uncertainty attribution analyzes that identify characteristics of cEEG associated with high AI model uncertainty. Together, these analyzes offer qualitative insight into when and why the hybrid system prioritizes novice input, without implying exhaustive coverage or guarantees of behavior.

**Hybrid decision maps across novice operating points.**

The hybrid system produces a continuous interval-level *novice-reliance weight* (denoted $w \in [0, 1]$), where larger values increase the contribution of the novice prediction and smaller values increase the contribution of the AI model's prediction. To interpret *why* the hybrid system behaves differently across novices, we visualize the learned decision surface over two clinically meaningful drivers that are available at inference time:

The hybrid system generates a weight (denoted $w \in [0, 1]$), where higher values increase the contribution of the novice, while lower values increase the contribution of the AI model. To interpret why the hybrid system behaves differently across novices, we visualize the decision surface over two clinically meaningful variables: (i) AI model's uncertainty, and (ii) cEEG complexity (higher complexity implies a more variable and artifact-prone signal). For each novice, we show:

- a **continuous decision map** (heatmap) over **AI model uncertainty** and **cEEG complexity**, where each cell reports the mean predicted weight for that region, and
- a **simplified 2-by-2 summary map** obtained by binarizing each axis of the continuous decision map into *AI model certain vs. uncertain* and *cEEG simple vs. complex*, where each cell reports the average predicted weight for that region.

These heatmaps allow us to reason about the hybrid system's policy in four interpretable regimes (AI model certain/uncertain by cEEG simple/complex), and to correlate that policy with each novice's error profile.

*Novice profiles.* Tables S9–S10 summarize the novice operating points. Novice N1 exhibits a *sensitivity-leaning but still specific* profile (high sensitivity with relatively strong positive predictive value and specificity), whereas novice N4 exhibits a *conservative* profile (very high specificity/PPV, but substantially lower sensitivity). These complementary behaviors create different opportunities for the hybrid system: N1 is particularly valuable when missed events are likely (recovering false negatives), while N4 is particularly valuable when false alarms are likely (suppressing false positives).

*Decision map for N1: sensitivity-leaning but clinically balanced.* Figure S2 shows that the hybrid system assigns low weights (trust AI) when the AI model is *certain*, and increases the weight (lean on the novice) as AI model uncertainty increases. This creates a clear gradient predominantly along the **AI-uncertainty**

Table S9: **Operating characteristics of novice N1 (sensitivity-leaning, balanced behavior).** Novice N1 demonstrates a high sensitivity while maintaining strong specificity and a positive predictive value, showcasing behavior that balances event detection and minimizes false alarms.

| Metric | Value |
|---|---|
| Accuracy | 0.99 |
| Positive Predictive Value (PPV) | 0.68 |
| Sensitivity | 0.83 |
| Specificity | 0.99 |
| True positives (TP) | 237 |
| False positives (FP) | 110 |
| True negatives (TN) | 11605 |
| False negatives (FN) | 48 |

Table S10: **Operating characteristics of novice N4 (high-specificity, conservative behavior).** Novice N4 demonstrates strict decision thresholds, which lead to high specificity and positive predictive value but lower sensitivity. This conservative approach minimizes false alarms but misses more true events.

| Metric | Value |
|---|---|
| Accuracy | 0.98 |
| Positive Predictive Value (PPV) | 0.72 |
| Sensitivity | 0.54 |
| Specificity | 0.99 |
| True positives (TP) | 153 |
| False positives (FP) | 60 |
| True negatives (TN) | 11655 |
| False negatives (FN) | 132 |

**axis**, consistent with the idea that the hybrid system uses uncertainty as a primary trigger to seek additional signal from the novice when the AI model is unreliable.

The 2×2 summary further reveals a second, novice-specific modulation by **cEEG complexity**: when the AI model is uncertain, the hybrid system weight is high in both EEG-simple and EEG-complex regimes, but is *especially elevated* in the EEG-complex regime. This behavior is aligned with the N1 profile: because N1 maintains strong sensitivity without an extreme false-positive burden, leaning on N1 is beneficial in hard (uncertain and/or complex) regimes where the AI model may miss subtle events or become unstable due to nonstationarities. In contrast, when the AI model is certain, the hybrid system remains AI-dominant, and only modestly increases weight with cEEG complexity—reflecting a conservative choice to avoid unnecessary novice-induced false alarms when the AI model already appears confident.

*Decision map for N4: conservative, high-specificity novice.* Figure S3 exhibits a qualitatively different policy. The most prominent pattern is an even cleaner gradient along the **AI-uncertainty axis**: the hybrid system increases $w$ from AI-certain to AI-uncertain conditions, indicating that the hybrid primarily consults N4 when the AI model is uncertain.

Crucially, relative to N1, the **EEG-complexity axis plays a weaker role** for N4: the 2×2 map shows similar weights for EEG-simple vs. EEG-complex within the AI-uncertain regime, with only a modest increase (if any) when complexity rises. This is a sensible adaptation to N4's conservative operating point: because N4 has excellent specificity and PPV but lower sensitivity, N4 is most useful as a *precision-preserving filter* when the AI model is unsure and at risk of false positives. However, in highly complex regimes, over-relying on a conservative novice could amplify false negatives (missed events), so the hybrid system avoids aggressively escalating $w$ purely due to EEG complexity. In other words, the hybrid system largely uses AI model uncertainty to decide *when* to consult N4, while limiting complexity-driven escalation that could otherwise suppress sensitivity.

*Contrasting N1 vs. N4: the same inputs, different novice-specific policies.* Taken together, Figures S2 and S3 illustrate the central mechanism: **the hybrid system learns novice-specific reliance rules over the same uncertainty/complexity space.** Both novices induce a dominant reliance gradient along AI model uncertainty (a shared, model-driven signal of reliability). Beyond that shared structure, the hybrid system adapts to the novice's operating point:

- For **N1** (high sensitivity with strong specificity), the hybrid system increases reliance not only when AI model uncertainty rises, but also more noticeably when cEEG complexity increases—especially in the AI-uncertain regime—reflecting that N1 can add value in difficult segments without a prohibitive false-alarm cost.

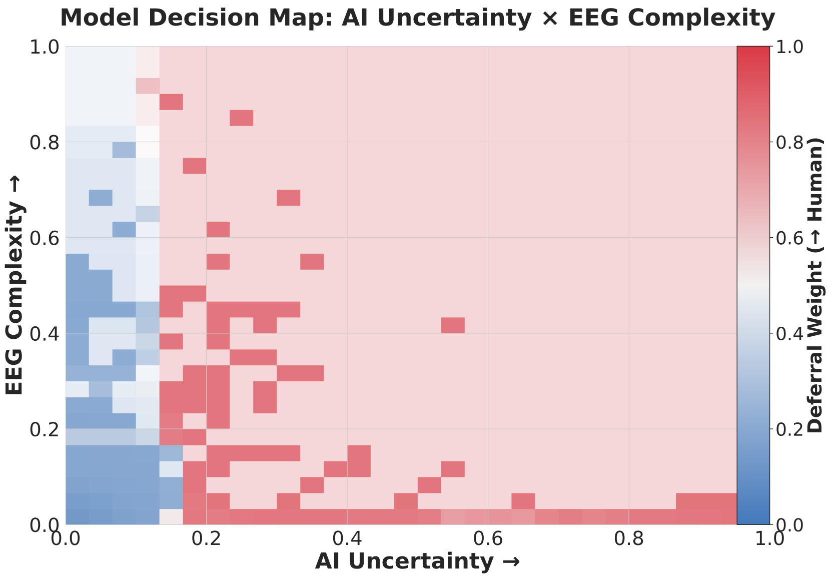

(a) **Continuous decision map** (AI model uncertainty × EEG complexity → $w$).

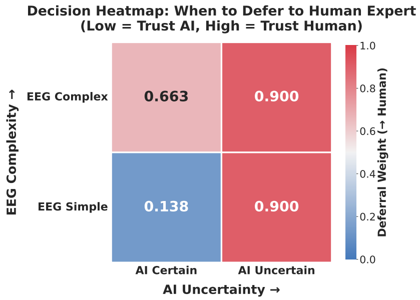

(b) **Coarsened 2×2 summary** (AI model certain/uncertain × EEG simple/complex).

Fig. S2: **Hybrid system decision maps for novice N1 (sensitivity-leaning, balanced behavior).** Heatmap color encodes the learned novice-reliance weight $w$, with warmer colors indicating greater reliance on the novice and cooler colors indicating greater reliance on the AI. Color transparency reflects data density, with darker regions indicating higher concentration of observed intervals (predominantly low cEEG complexity and low-to-moderate AI model uncertainty), and progressively lighter regions indicating sparser support toward higher complexity and uncertainty. The dominant structure is a strong gradient along the AI model uncertainty axis, with deferral increasing as the AI model becomes less confident. A secondary, but consistent, modulation is observed along cEEG complexity, particularly in the AI-uncertain regime, indicating increased reliance on N1 in harder conditions where complex signals coincide with AI model uncertainty and N1's sensitivity-leaning profile is most beneficial.

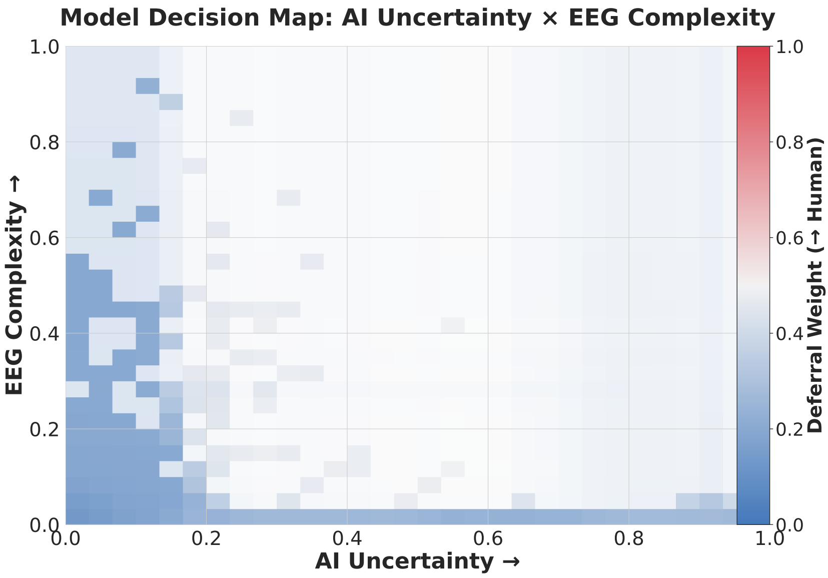

(a) **Continuous decision map** (AI model uncertainty $\times$ EEG complexity $\rightarrow$ $w$).

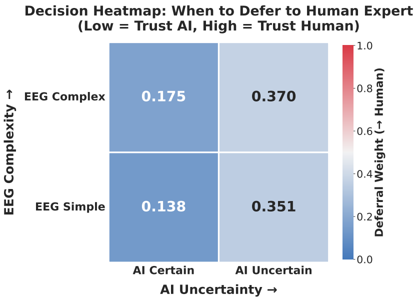

(b) **Coarsened 2×2 summary** (AI model certain/uncertain $\times$ EEG simple/complex).

Fig. S3: **Hybrid system decision maps for novice N4 (conservative, precision-preserving behavior).** Heatmap color encodes the learned novice-reliance weight $w$, while color transparency reflects data density, highlighting that most observations lie in the low-complexity, low-to-moderate uncertainty region and that the upper-right portion of the map is sparsely populated. Deferral exhibits a strong monotone dependence on AI model uncertainty, with increasing reliance on N4 as the AI model becomes uncertain. In contrast to N1, modulation by cEEG complexity is comparatively weak, consistent with N4's conservative operating profile (high PPV and specificity, lower sensitivity) and the limited evidence for true positive events in high-complexity regimes. Accordingly, the hybrid system primarily leverages N4 to suppress false alarms under AI model uncertainty rather than escalating deferral solely based on signal complexity.

– For **N4** (high PPV/specificity, lower sensitivity), the hybrid system relies on N4 primarily as a function of AI model uncertainty and keeps the complexity effect modest, reflecting a calibrated trade-off: consult a conservative novice when the AI model is uncertain, but avoid over-weighting the novice in complex regimes where missed events would be costly.

Taken together, Figures S2 and S3 illustrate a unifying mechanism underlying hybrid system performance: **AI model uncertainty governs *when* the hybrid system consults a novice, while novice-specific error profiles govern *how* that consultation is carried out.** Across novices, the dominant and consistent structure is a monotone reliance gradient along the AI-uncertainty axis, indicating that the hybrid system first decides whether the AI model itself is reliable before modulating reliance based on cEEG complexity or novice characteristics.

Beyond this shared structure, the hybrid system adapts its behavior to each novice's operating point. For sensitivity-leaning novices such as N1, reliance increases more strongly in complex regimes once uncertainty is high, reflecting the novice's ability to recover missed events without excessive false alarms. For conservative novices such as N4, reliance remains tightly coupled to AI model uncertainty while limiting complexity-driven escalation, reflecting a calibrated use of the novice as a precision-preserving filter.

This contrast provides an interpretable explanation for how a single hybrid system framework can consistently improve performance across heterogeneous novices: it exploits structured disagreement by learning where each novice is most helpful, rather than treating all novice input as uniformly beneficial.

#### Uncertainty explainability: methods and results

Because the proposed hybrid system adaptively relies on novice input primarily when the AI model is uncertain, understanding the *drivers of AI uncertainty* is critical for interpreting the behavior of the deferral mechanism. In this subsection, we perform a SHAP-based explainability study to identify which cEEG features most strongly contribute to elevated AI uncertainty.

*Overview of the Uncertainty Explainability Pipeline.* The uncertainty explainability analysis proceeds in three stages: feature aggregation, uncertainty quantification, SHAP analysis

*Feature aggregation:*

Starting from the full 111-feature cEEG representation, we construct a reduced, interpretable feature set by averaging left–right hemispheric pairs within each frequency band and montage (e.g., Delta F3–P3 and Delta F4–P4 are averaged into a single Delta F–P feature). This yields a compact set of 16 features corresponding to four canonical cEEG frequency bands (delta, theta, alpha, beta) across four montages (F–P, P–O, F–T, T–O). This aggregation preserves physiologically meaningful structure while reducing redundancy and noise.

*Uncertainty Quantification:*

An Extra Trees classifier is trained to predict the clinical outcome using these 16 aggregated cEEG features. For each cEEG interval, we compute predictive uncertainty as the entropy of the model's predicted class probabilities. Entropy values near zero correspond to confident predictions (probability near 0 or 1), while higher entropy values indicate ambiguity, with maximum uncertainty occurring when predicted probabilities approach 0.5.

*SHAP analysis of AI uncertainty:*

To identify which cEEG features drive AI uncertainty, we model predictive entropy as a function of the same 16 aggregated cEEG features and apply SHAP to interpret feature contributions. This analysis answers the question: *which cEEG characteristics most strongly increase or decrease the AI model's uncertainty?* By computing SHAP values with respect to uncertainty itself—rather than the clinical label—the results directly characterize the signals that trigger the hybrid system's deferral behavior.

*Results: Dominance of Alpha and Theta Band Features.* Figure S4 summarizes the mean absolute SHAP values for all 16 features, grouped by frequency band. Two clear trends emerge.

First, alpha-band features exhibit the largest contributions to AI uncertainty across multiple montages, particularly in fronto–parietal and fronto–temporal regions. Second, theta-band features also contribute substantially to uncertainty, whereas delta and beta bands play a comparatively minor role.

Why Alpha and Theta Dominance Is Expected? The prominence of alpha and theta activity as drivers of AI uncertainty is physiologically and clinically plausible in the context of intraoperative cEEG monitoring.

Alpha and theta rhythms occupy an intermediate regime between highly pathological and clearly normal cEEG patterns. During anesthesia and cerebral hypoperfusion, these bands are particularly sensitive to changes in arousal, anesthetic depth, cortical deafferentation, and early ischemic stress. Consequently, alpha and theta features often reflect *transitional or mixed states* rather than unequivocal pathology.

From an ML perspective, such transitional regimes naturally induce predictive ambiguity. Unlike delta-band suppression or burst attenuation—which often cor-

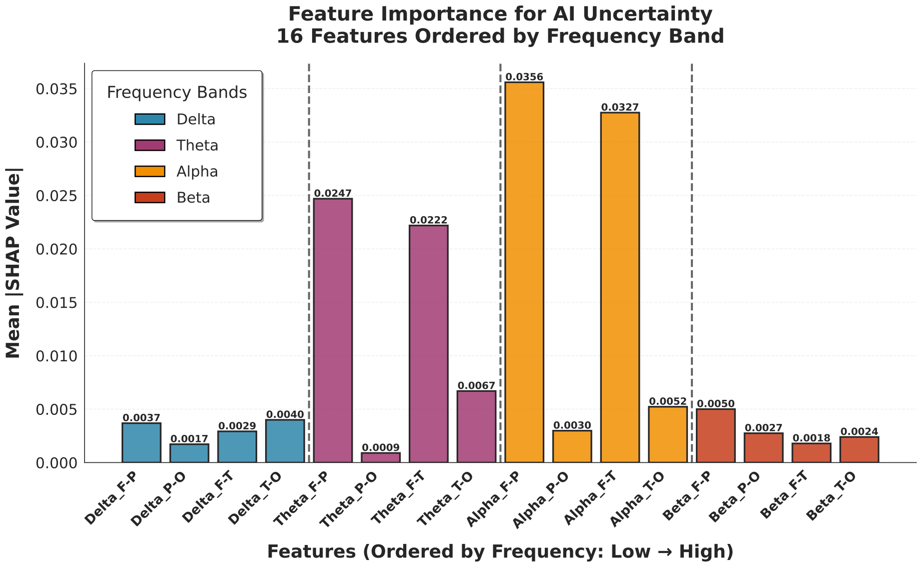

Fig. S4: **SHAP feature importance for AI predictive uncertainty.** Mean absolute SHAP values from an entropy-based uncertainty model using 16 aggregated cEEG features (4 frequency bands $\times$ 4 montages). Features are ordered by frequency band from low to high. Alpha- and theta-band features dominate uncertainty contributions, indicating that intermediate-frequency cEEG activity is the primary driver of AI predictive ambiguity.

respond to more stereotyped and easily separable cEEG patterns—alpha and theta activity can arise from multiple physiological processes with overlapping spectral signatures. As a result, similar alpha/theta configurations may correspond to both positive and negative outcomes in the training data, leading the AI model to assign intermediate probabilities and thus higher entropy.

Importantly, this behavior aligns with the intended role of the hybrid system. Intervals with ambiguous alpha- and theta-dominant activity are precisely those where the AI model is least reliable and where human pattern recognition can add value. The finding that alpha and theta features drive AI uncertainty supports the mechanistic validity of the deferral policy: the hybrid system does not defer arbitrarily, but in physiologically meaningful situations where uncertainty is inherently high.

From the perspective of the hybrid system, these same regimes correspond to intervals in which novice input is most likely to add value. Because the hybrid defers to the novice primarily when AI uncertainty is elevated, the dominance of alpha- and theta-band features as drivers of uncertainty implicitly identifies the cEEG conditions under which novice judgment is preferentially leveraged. In other words, novice expertise is not applied uniformly across all cEEG patterns, but is selectively recruited in intermediate-frequency regimes where the AI's decision boundary is intrinsically ambiguous.

*Implications for Hybrid Decision-Making.* This analysis demonstrates that AI uncertainty is not driven by noise or artifact-dominated features, but instead arises from clinically meaningful cEEG dynamics associated with intermediate-frequency activity. Because AI uncertainty serves as the primary trigger for novice reliance in the hybrid novice-AI system, these results also clarify *where novice input is most beneficial*: specifically, in alpha- and theta-dominant regimes that reflect transitional or mixed physiological states.

Together, the decision maps and uncertainty attribution results provide a mechanistic account of the hybrid policy under typical operating conditions. In the next section, we present representative intraoperative case studies to qualitatively illustrate how this policy manifests in real cEEG recordings affected by common artifacts and physiologic confounds, offering additional insight into hybrid behavior in challenging and edge-case scenarios.

#### S7 Robustness to cEEG Artifacts

Intraoperative cEEG signals are frequently contaminated by artifacts arising from surgical manipulation, anesthesia, and hardware instability, which can compromise the interpretation of ischemia. We discuss these common artifacts and detail the robustness of the hybrid novice–AI system against them.

**Electrocautery**

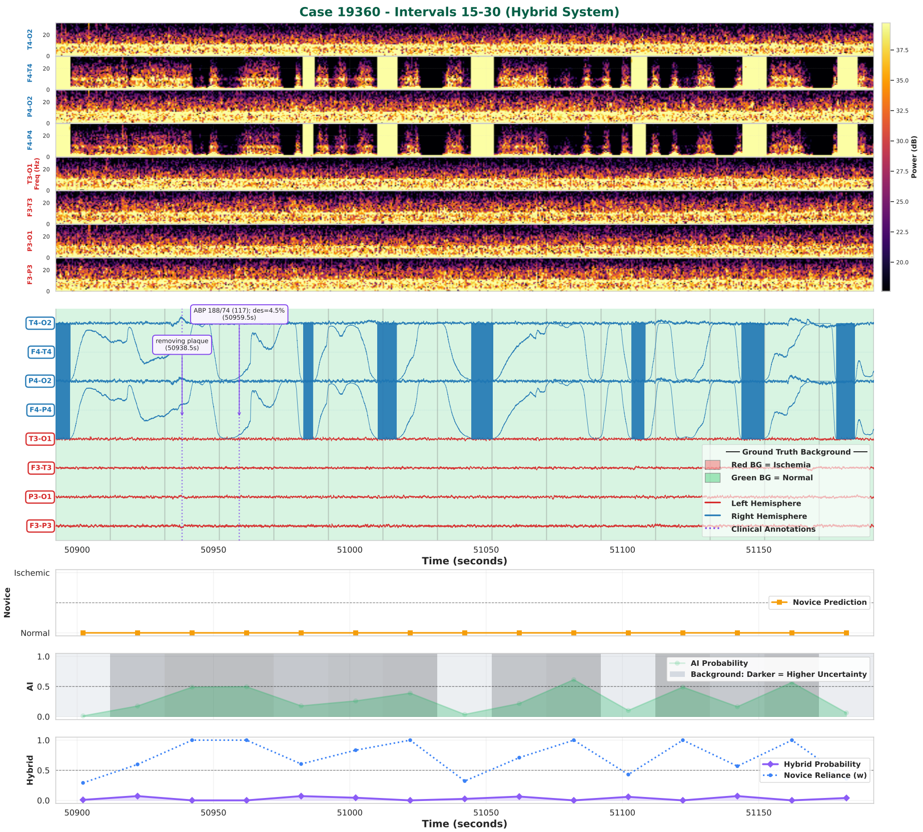

Fig. S5: **Electrocautery artifact.** An example illustrating the robustness of the hybrid system to electrocautery-induced artifacts. Repeated electrocautery generates high-amplitude noise in the right-hemispheric channels F4–T4 and F4–P4, which is evident in both the raw cEEG and the density spectral array (DSA).

Electrocautery often generates broadband, high-amplitude noise that can obscure underlying neural activity. In the example shown in Figure S5, multiple episodes of electrocautery result in corruption of signals at the channel level. The AI model exhibits increased uncertainty and unstable ischemia probabilities, whereas the novice accurately identifies the artifacts. The hybrid system relies more on the novice when the AI's uncertainty is high, thereby preventing false positives.

#### Electrode Pop

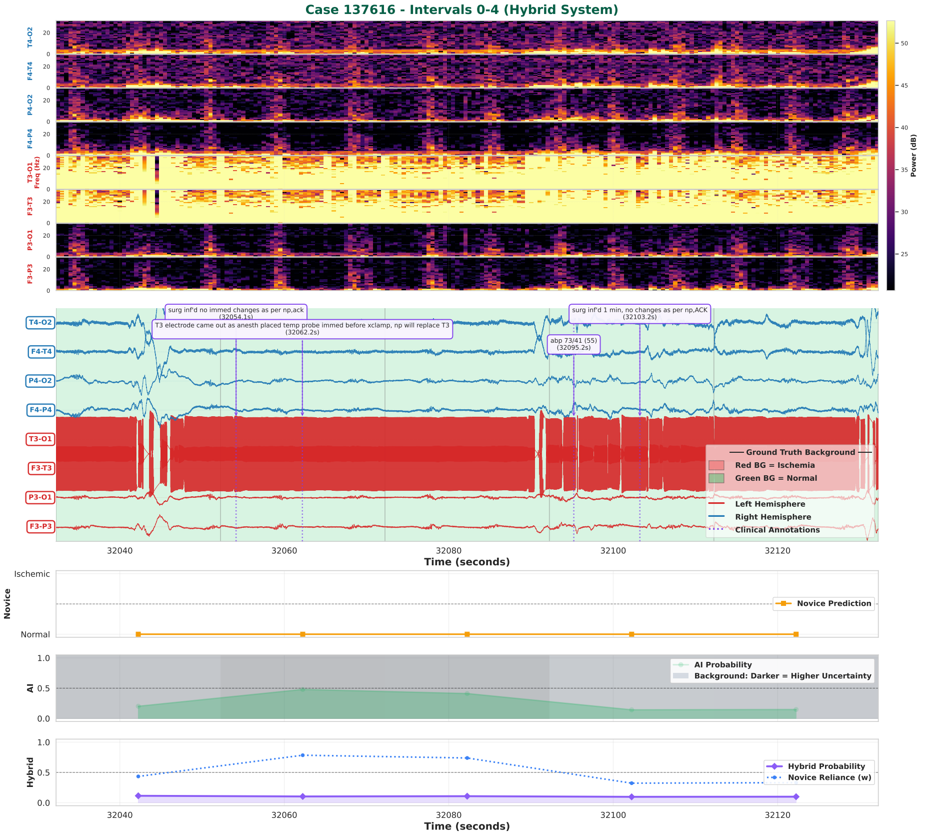

Fig. S6: **Electrode pop artifact.** An example illustrating the robustness of the hybrid system to electrode pop artifacts. Dislodgement of the T3 electrode results in high-amplitude artifacts in the left-hemispheric channels F3–T3 and T3–O1, visible as high-frequency signals in the raw cEEG and uniformly elevated broadband power in the DSA.

Electrode pop is a nonphysiological, high-amplitude, short-duration artifact that appears as a sharp, sudden voltage spike caused by a loose or poorly applied electrode. In the example shown in Figure S6, the T3 electrode pop produces pronounced artifacts in multiple channels. The AI model exhibits increased uncertainty, whereas the novice accurately identifies the artifacts. The hybrid system relies more on the novice when the AI's uncertainty is high, thereby preventing false positives.

#### Anesthesia

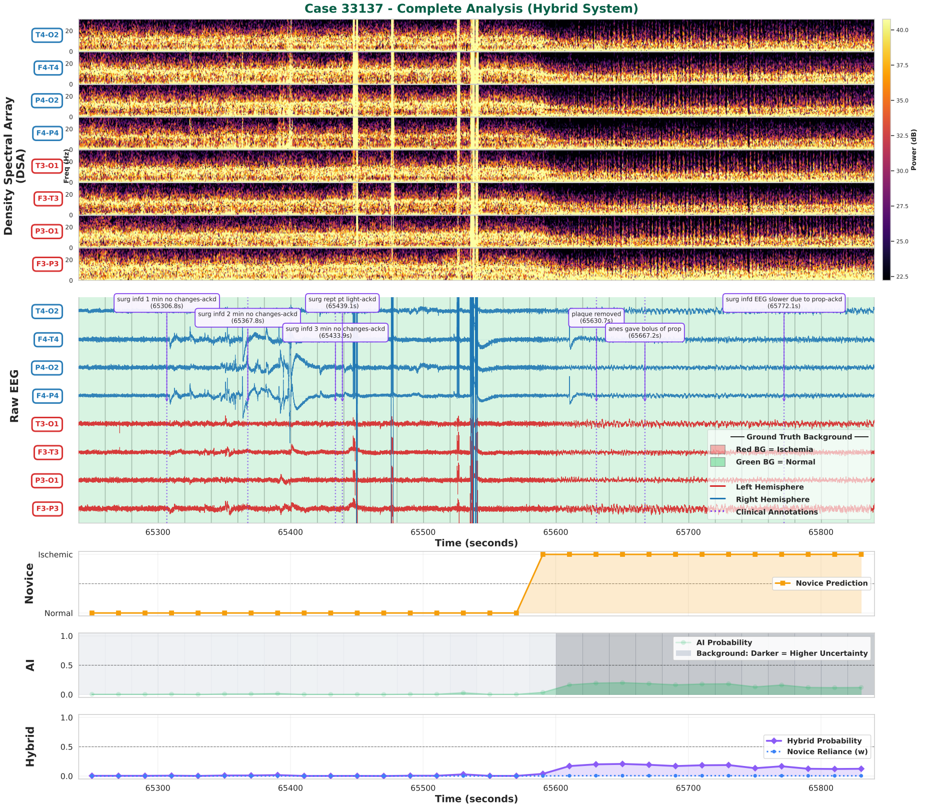

Fig. S7: **Anesthetic-induced artificat.** An example illustrating the robustness of the hybrid system in anesthetic-induced cEEG slowing. The anesthetic propofol produces diffuse cEEG slowing and reduced spectral power in the DSA across all channels, resembling ischemia.

Certain anesthetic agents, such as propofol, induce diffuse cEEG slowing and power attenuation that can mimic ischemia. In the example shown in Figure S7, there is slowing across all channels following propofol administration. The novice incorrectly identifies this as ischemia, whereas the AI model is able to distinguish anesthetic-induced slowing from ischemia with certainty using spectral power features. The hybrid system correspondingly places greater weight on the stable, low-uncertainty AI prediction, thereby preventing false positives.

#### Benign Interhemispheric Asymmetry

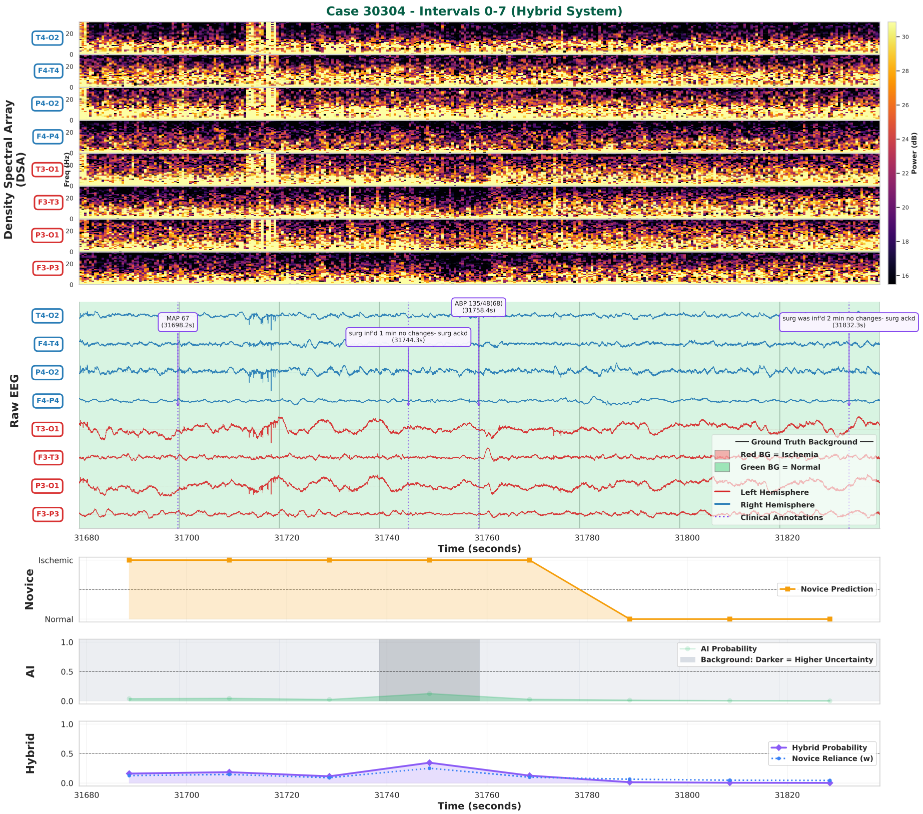

Fig. S8: **Benign interhemispheric asymmetry.** An example illustrating the robustness of the hybrid system in the presence of benign interhemispheric asymmetry. Mild, stable asymmetry in amplitude and spectral power across all channels.

In some patients, there is asymmetry in intraoperative cEEG between hemispheres that is benign and not caused by ischemia, such as the example shown in Figure S8. The novice incorrectly identifies the asymmetry as ischemia despite the absence of progressive temporal or spectral changes. The AI model correctly identifies the pattern as not ischemic with low uncertainty, which allows the hybrid system to defer to the AI model and avoid false positives.

#### Joint Failure of Novice and AI

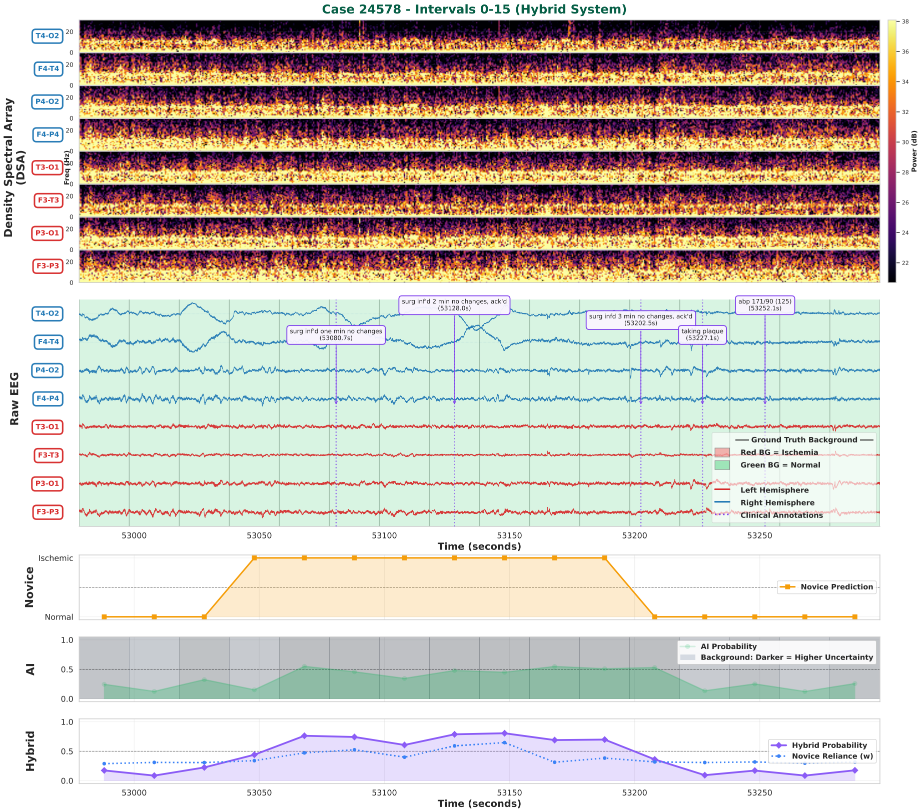

Fig. S9: **Limitation.** An example illustrating the limitation of the hybrid system. When both the AI model and the novice are incorrect, the hybrid system combines their errors, yielding a false positive.

In the example illustrated in Figure S9, the hybrid system generates a false positive because both the AI model and the novice are incorrect. This situation highlights that the hybrid system is not infallible; its advantages are realized only when at least one input is accurate. This implies that the hybrid system can be improved in the future by expert escalation, abstention mechanisms, or additional contextual inputs.

Table S11: **Calibration analysis comparing hybrid and AI systems on dataset-1.** Values are reported as mean ± standard deviation across 200 paired bootstrap replicates. Lower values indicate better calibration for all metrics. P-values were computed using a paired bootstrap procedure and represent the proportion of replicates in which the AI system outperformed the hybrid system.

| Metric | AI (mean ± SD) | Hybrid (mean ± SD) |
|---|---|---|
| Brier score | $0.016 \pm 0.001$ | $0.013 \pm 0.001$ ($p < 0.001$) |
| Log loss | $0.057 \pm 0.003$ | $0.049 \pm 0.006$ ($p < 0.025$) |
| Expected calibration error (ECE) | $0.002 \pm 0.0006$ | $0.0018 \pm 0.0007$ ($p = 0.41$) |
| Maximum calibration error (MCE) | $0.010 \pm 0.005$ | $0.007 \pm 0.005$ ($p = 0.35$) |

#### S8 Calibration Analysis

In addition to the calibration results based on proper scoring rules provided in the main text, we evaluated bin-based calibration metrics, including expected calibration error (ECE) and maximum calibration error (MCE). Calibration performance was evaluated using a paired bootstrap procedure with 200 bootstrap replicates, comparing per-replicate differences between the hybrid and AI systems on the same resampled data.

As shown in Table S11, the hybrid system achieved a lower mean Brier score and log-loss than the AI model, with consistent superiority across bootstrap replicates, indicating modest improvements in probabilistic accuracy under proper scoring rules. For ECE and MCE, the hybrid system also exhibited lower mean values, suggesting improved average calibration; however, these differences were not statistically significant (p = 0.41 for ECE and p = 0.35 for MCE), reflecting higher variability inherent to bin-based calibration metrics.

Taken together, these results indicate that the hybrid approach preserves the AI model's calibration and may yield small directional improvements in bin-based calibration, while demonstrating more robust and consistent gains under proper scoring rules. Consistent with established practice in probabilistic prediction modeling, we emphasize proper scoring rules—specifically, Brier score and log loss—as primary measures of calibration performance. Both metrics are strictly proper scoring rules that provide threshold-free, sample-efficient summaries of probabilistic accuracy and jointly capture calibration and sharpness (7; 24; 18).

Bin-based calibration metrics such as ECE and MCE are widely used as diagnostic tools, particularly in the ML literature (9); however, their empirical estimates are known to be sensitive to binning choices and can exhibit bias and variance, especially in finite samples (14; 16). For this reason, we report ECE and MCE in the Supplementary material for completeness, while relying on Brier score and log loss as the primary calibration metrics in the main analysis.

### S9 Robustness to the Choice of AI Model

In the main manuscript, we report our primary results using a single strong AI model (Extra Trees; ET) paired with novices, in order to keep the presentation focused and avoid conflating algorithmic contributions with model-selection effects. Here, we evaluate whether the core conclusions remain consistent when the hybrid system is instantiated with alternative AI models. Specifically, we ask two questions: (i) does the hybrid reliably improve over the underlying AI model in discrimination performance, and (ii) does the hybrid continue to satisfy the expert-level non-inferiority (NI) criterion even when paired with weaker or qualitatively different AI predictors.

#### Choice of AUPRC in a rare-event setting

We report both the area under the precision–recall curve (AUPRC) and the area under the receiver operating characteristic curve (AUROC). Because ischemic events are rare in our dataset (prevalence ≈2.4%), resulting in class imbalance, AUPRC can provide complementary insight into performance on the minority (ischemic) class. In rare-event settings, AUROC may remain high even when precision is modest, as negative samples dominate (20; 4; 23). By contrast, AUPRC directly summarizes the trade-off between precision (positive predictive value) and recall (sensitivity), reflecting how often an alarm corresponds to a true ischemic event (20; 15).

Recent theoretical work further clarifies this distinction. (11) showed that under severe class imbalance ($a = N^-/N^+ \gg 1$), AUROC and AUPRC can diverge substantially: a fixed AUROC value may correspond to a wide range of AUPRC values. In our setting ($a \approx 40.7$), AUROC values near 0.9 could still correspond to a broad span of precision–recall performance. Accordingly, AUPRC is particularly informative for interpreting minority-class behavior and is considered alongside AUROC in our analysis.

#### Robustness across AI models

We evaluate hybrid performance across a diverse set of AI models, including Random Forest (RF; (2)), Extra Trees (ET; (6)), Light Gradient Boosting Machine (LightGBM; (10)), Extreme Gradient Boosting (XGBoost; (3)), Gradient Boosting Machines (GradBoost; (5)), multilayer perceptrons (MLP; (19; 17; 13)), FT-Transformer (FTT; (8)), and TabNet ((1)). These models span bagging-based ensembles, boosting methods, and deep learning architectures for tabular data.

Across all evaluated AI models, each corresponding hybrid system yields substantial improvements in AUPRC relative to its underlying AI model (Table S12), with absolute gains ranging from approximately 15% to 40%. The magnitude of improvement depends on the AI model performance: stronger models such

Table S12: **Hybrid novice-AI systems consistently outperform both the AI model and the novices across diverse AI models.** Across all evaluated AI models, each corresponding hybrid system—built on top of that same AI model—yields large and consistent improvements in discrimination, increasing AUPRC by approximately 15–40% relative to its underlying AI model. Despite substantial heterogeneity in AI architectures and performance, every hybrid system satisfies the expert non-inferiority criterion, whereas none of the novices do. These results show that the hybrid system consistently improves the specific AI model it is paired with, achieving expert-level behavior while substantially improving precision–recall performance in low-prevalence settings. Reported values are mean ± SD AUPRC. AUPRC denotes the absolute improvement of the hybrid over its underlying AI model.

| **AI model** | **AI AUPRC** | **Hybrid AUPRC** | **$\Delta$ AUPRC** | **Novice meets NI?** | **Hybrid meets NI?** |
|---|---|---|---|---|---|
| ET | $0.54 \pm 0.03$ | **0.72** $\pm$ 0.05 | +0.18 | ✗ | ✓ |
| RF | $0.53 \pm 0.03$ | **0.68** $\pm$ 0.03 | +0.15 | ✗ | ✓ |
| LGBM | $0.47 \pm 0.03$ | **0.69** $\pm$ 0.05 | +0.22 | ✗ | ✓ |
| XGB | $0.46 \pm 0.03$ | **0.74** $\pm$ 0.08 | +0.28 | ✗ | ✓ |
| TabNet | $0.35 \pm 0.04$ | **0.71** $\pm$ 0.07 | +0.36 | ✗ | ✓ |
| MLP | $0.34 \pm 0.04$ | **0.62** $\pm$ 0.07 | +0.28 | ✗ | ✓ |
| FTT | $0.32 \pm 0.03$ | **0.66** $\pm$ 0.08 | +0.34 | ✗ | ✓ |
| GradBoost | $0.26 \pm 0.03$ | **0.67** $\pm$ 0.09 | +0.41 | ✗ | ✓ |

✓: passes expert non-inferiority test; ✗: does not pass.

as Random Forest and Extra Trees exhibit moderate absolute gains, whereas weaker or less well-calibrated models such as TabNet and Gradient Boosting exhibit larger improvements. This monotonic pattern is expected in a complementary decision system: when the AI model is less reliable on subsets of cEEG intervals, the deferral mechanism increases reliance on the novice in precisely those regimes, translating into larger net gains in precision–recall performance.

Importantly, while none of the novices satisfy the expert non-inferiority criterion under our predefined statistical testing procedure, all hybrid instantiations do. Thus, the hybrid's expert-level behavior is not an artifact of selecting a single AI model (ET), but rather appears to be a stable property of the proposed integration strategy across heterogeneous AI models. Taken together, these results support the conclusion that the hybrid novice-AI system robustly adapts to diverse AI model choices while delivering expert-level performance and substantial improvements in precision–recall behavior in a low-prevalence, high-imbalance clinical setting.

Taken together, these results demonstrate that the proposed hybrid novice-AI system is robust to the choice of AI model: across a wide range of model families and performance levels, the hybrid consistently improves discrimination performance by 15–40% in AUPRC and achieves expert-level behavior as assessed by the non-inferiority test. This robustness indicates that the reported gains are not driven by a particular AI architecture, but reflect a general property of the hybrid integration strategy in low-prevalence, high-imbalance clinical settings.

### S10 Why Hybrid Systems Work: Decision-Level Diversity Between AI Models and Novices

##### Why does the hybrid work reliably?

While the main and supplementary results demonstrate that the proposed hybrid system consistently improves discrimination performance and achieves expert-level behavior across diverse AI models, these findings naturally raise a deeper question: *why* does the hybrid work so reliably? In this section, we provide a mechanistic analysis of the hybrid novice-AI system by examining decision-level agreement and disagreement both across AI models and between AI models and novices. Our central thesis is that the hybrid system succeeds because it operates in a regime of *structured, heterogeneous disagreement*, which the deferral mechanism can exploit adaptively on a per-interval basis.

##### Decision-level diversity among AI models (Cohen's $\kappa$)

We first examined decision-level similarity across AI models by computing pairwise Cohen's $\kappa$ between thresholded binary predictions. This analysis complements standard performance metrics such as AUPRC: whereas AUPRC quantifies discriminative ability under class imbalance, Cohen's $\kappa$ characterizes whether different models tend to raise alarms on the same cEEG intervals beyond chance.

Figure S10 visualizes these pairwise agreements after ordering models by architectural similarity and decision coherence. The resulting structure is highly non-random. Tree-based ensembles trained via bagging (Extra Trees, Random Forest) form a tight high-agreement block, reflecting near-identical decision behavior on many cEEG intervals. Closely related gradient-boosted tree models (XGBoost, LightGBM, Gradient Boosting) form a second coherent cluster with strong within-group agreement and moderate agreement with the bagged ensemble block.

In contrast, neural-network–based tabular models exhibit systematically different behavior. The FT-Transformer (FTT) occupies an intermediate regime: its agreement with tree-based models is consistently lower than intra-tree agreement (e.g., Random Forest - Extra Trees or LGBM–XGB), yet substantially higher than that of a generic multilayer perceptron (MLP). TabNet similarly shows intermediate agreement, indicating a distinct inductive bias that partially overlaps—but does not coincide—with tree-based decision logic. The MLP stands

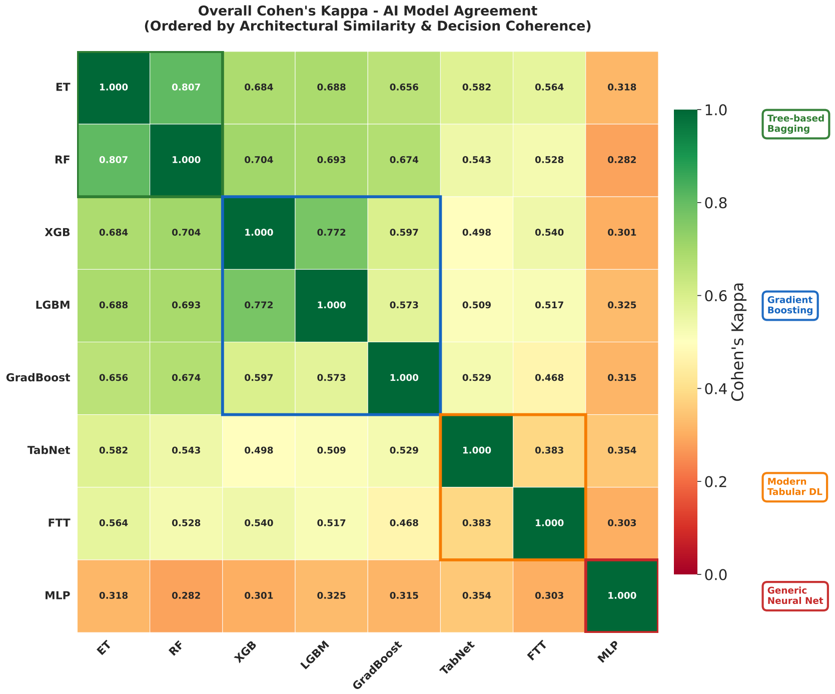

Fig. S10: **Decision-level similarity among AI models.** Pairwise Cohen's $\kappa$ computed between thresholded binary predictions from each AI models, ordered by architectural similarity and decision coherence. Higher values indicate more similar alarm decisions beyond chance. Clear block structure emerges: bagged tree ensembles and gradient-boosted trees exhibit high within-group agreement, whereas tabular deep learning models (FTT, TabNet) and especially MLP show progressively lower agreement, reflecting distinct inductive biases and decision patterns.

apart with uniformly low agreement across nearly all models, forming a visually and quantitatively isolated block.

Taken together, the block-diagonal structure in Fig. S10 confirms that the evaluated AI models span a spectrum of qualitatively distinct decision behaviors rather than constituting a homogeneous set. As a result, robust hybrid performance across all models is not a trivial consequence of shared errors or redundant decision logic.

**Decision-level disagreement between AI models and novices.**

We next analyzed agreement between AI models and novices using Cohen's $\kappa$, again computed on thresholded binary decisions. Overall, AI–novice agreement is modest, with most $\kappa$ values falling in the "slight" to "fair" range, indicating that AI models and novices frequently disagree on which cEEG intervals warrant alarms.

Averaged across novices, stronger tree-based models such as Random Forest (RF), LightGBM (LGBM), Extra Trees (ET), and XGBoost (XGB) exhibit the highest agreement with novice decisions (average $\kappa \approx 0.30$). The FT-Transformer (FTT) shows moderate agreement ($\kappa \approx 0.25$), exceeding that of a standard multilayer perceptron (MLP) but remaining lower than tree ensembles. In contrast, MLP exhibits substantially lower agreement with novice decisions ($\kappa \approx 0.13$), while TabNet and Gradient Boosting occupy an intermediate regime. These trends are summarized in Table S13 (left).

Agreement also varies systematically across novices. When averaged across AI models (Table S13, right), three novices (N1, N2, and N3) display comparable levels of agreement with AI models ($\kappa \approx 0.27$), whereas one novice (N4) exhibits consistently lower agreement ($\kappa \approx 0.20$). This confirms that novices are not interchangeable and differ meaningfully in how they interpret cEEG intervals relative to algorithmic predictions.

Table S13: **Decision-level agreement between AI models and novices.** Average Cohen's $\kappa$ values summarizing agreement (left) across all novices for each AI model, and (right) across all AI models for each novice.

| **AI model** | **Avg. Cohen's $\kappa$** | **Novice** | **Avg. Cohen's $\kappa$** |
|---|---|---|---|
| RF | 0.32 | N1 | 0.27 |
| LGBM | 0.31 | N2 | 0.28 |
| ET | 0.31 | N3 | 0.27 |
| XGB | 0.30 | N4 | 0.20 |
| FTT | 0.25 | | |
| GradBoost | 0.24 | | |
| TabNet | 0.20 | | |
| MLP | 0.13 | | |

**Implications for hybrid decision-making.**

Crucially, low Cohen's $\kappa$ should not be interpreted as poor performance by either AI models or novices. Cohen's $\kappa$ measures agreement, not correctness. Instead, the observed disagreement indicates that AI models and novices tend to err on different subsets of cEEG intervals.

This structured, complementary disagreement—evident both across AI architectures and between AI and novices—is precisely the regime in which hybrid and learning-to-defer systems are expected to provide the greatest benefit. Consistent with this interpretation, even AI models that show low agreement with novices (e.g., MLP and TabNet) yield substantial gains in AUPRC when integrated into the hybrid system and satisfy the expert non-inferiority criterion.

In summary, the hybrid system does not rely on consensus between AI models and novices, nor on selecting a particular AI inductive bias. Instead, it exploits decision-level diversity and human–AI heterogeneity to adaptively arbitrate between competing predictions on a per-interval basis. The clear block structure in Fig. S10 provides mechanistic evidence that such heterogeneity is both substantial and systematic, explaining why the hybrid system performs robustly across models, novices, and clinical conditions.

Overall, the Supplementary analyses show that the hybrid system works well because the AI and the novice make different kinds of mistakes. By combining their outputs at each interval, the system can correct these errors. The strong performance does not come from agreement or from relying on the AI alone.

#### Code and Data Availability

The code developed in this study will be made publicly available for non-commercial academic use at:

`https://github.com/batmanlab/EEG-Hybrid-AI-Systems`.

The repository will include code for feature extraction from cEEG signals, for training AI models, and for implementing the hybrid novice–AI system. Because of patient privacy and data-sharing restrictions, we cannot publicly release raw cEEG recordings. However, we provide sufficient methodological details to enable replication of our analyses using appropriately approved cEEG datasets.
